# Ambient Temperature and Dengue Hospitalisation in Brazil: A 10-year period two-stage case-design time series analysis

**DOI:** 10.1101/2022.04.05.22273394

**Authors:** Rafael Lopes, Xavier Basagaña, Leonardo S. L. Bastos, Fernando A. Bozza, Otavio T. Ranzani

## Abstract

Climate factors are known to influence seasonal patterns of dengue transmission. However, little is known about the effect of extremes of temperature on the severity of dengue infection, such as hospital admission. We aimed to quantify the effect of ambient temperature on dengue hospitalisation risk in Brazil. We retrieved daily dengue hospitalisation counts by each of 5,565 municipalities across the 27 states of Brazil from 1st of January 2010 to 31st of December 2019, from the Brazilian Public Hospital Admission System (“SIH”). We obtained average daily ambient temperature for each municipality from the ERA5-land product reanalysis. We combined distributed lag non-linear models with time stratified design model framework to pool a relative risk (RR) estimate for dose-response and lag-response structures for the association of temperature and Dengue hospitalisation. We estimate the overall dengue hospitalisation RR for the whole country as well for each of the five macro-regions by meta-analysing state level estimates. 579,703 hospital admissions due to dengue occurred over the 10 years’ period of 2010 to 2019. We observed a positive association between high temperatures and high risk of hospitalisation for Brazil. The overall RR for dengue hospitalisation was 2.185 (95% CI 1.457- 3.276) at the 50th percentile of temperature and 2.385 (95% CI 1.556-3.655) at 95th percentile of temperature for Brazil. We also found lag effects of heat on hospitalisation, particularly an immediate augmented risk to hospitalisation, both at the 50th percentile and 95th. High temperatures are associated with an increase in the risk of hospitalisation by dengue. These findings may guide preparedness and mitigation policies during dengue season outbreaks, particularly on the health-care demand.

## 1. Introduction

Dengue fever has been a major global seasonal endemic disease, present largely in the tropics, with an estimated burden over ∼390 million infections per year (Bhatt et al., 2013; Guzman and Harris, 2015). Approximately one quarter of these infections manifest as clinical or subclinical disease (Bhatt et al., 2013). Brazil is one of the most affected countries by dengue, that has caused over ∼20.9 millions cases of infections in the last 20 years, since the compulsory universal notification to the Brazilian health systems (Godói et al., 2018; Lowe et al., 2021). Additionally, it is estimated that the Brazilian Universal Health system (SUS) spent more than USD 159 millions in the treatment and assistance to dengue cases and USD 10 million on severe dengue between 2000 and 2015 (Godói et al., 2018). The impressive burden of dengue in Brazil is also observed in several others low-income and middle-income countries (Suaya et al., 2009).

Dengue incidence is influenced by climate variables, particularly with temperature and rainfall, due to mosquito life cycle and human behaviour changes, resulting in increased transmission risk (Bhatt et al., 2013; Campbell et al., 2015; Colón-González et al., 2021; Lee et al., 2021; Lowe et al., 2021; Suaya et al., 2009; Wibawa et al., 2024). A systematic review and meta-analysis published in 2023 pooled 106 studies evaluating the association between high temperature and heatwaves with dengue incidence (Damtew et al., 2023). There was sufficient evidence to show increased relative risk for high temperatures (pooled RR 1.13 (95% CI, 1.11-1.16, for 1°C increase in temperature), however limited evidence for heatwave events (pooled RR 1.08, 95% CI, 0.95-1.23). The majority of articles used a monthly time resolution of the exposure, usually exploring lag effects up to one year. The magnitude of the association reported seemed to depend on the climate zone, showing a greater incidence risk in the tropical monsoon and humid subtropical climate zones (Damtew et al., 2023). In this scenario, projections show that dengue incidence, as well as other mosquito-borne diseases such as Zika, can have increased epidemic potential with the global rising temperature due to climate change (Colón-González et al., 2021; Van Wyk et al., 2023).

Dengue fever can present with increasing severity, requiring hospital admission and the support of vital organs (Paz-Bailey et al., 2024). The main mechanisms involved in dengue severity comprises dehydration and coagulation disorders (Burattini et al., 2016; Paz-Bailey et al., 2024; Werneck et al., 2018), both conditions which can be aggravated by ambient temperature (Kenney et al., 2014; Schneider et al., 2017). Although the literature is clear regarding the association between temperature and dengue transmission, there is a lack of evidence regarding the association between ambient temperature and dengue severity. Based on the reported association of short-term extremes of temperature and all-cause hospitalizations (Martínez-Solanas and Basagaña, 2019; Pudpong and Hajat, 2011; Vaidyanathan et al., 2019), we hypothesised that short-term ambient temperature has a positive association with the risk of hospitalisation due to dengue. We aimed to evaluate the association between ambient temperature and dengue hospitalisation in Brazil, analysing the period from 2010 to 2019.

## 2. Methods

### 2.1 Study design

We conducted a two-stage case-design time series analysis to evaluate the association between ambient temperature and dengue hospitalisation in Brazil. The unit of analysis was municipality of residence.

### 2.2 Study area

Brazil is located in South America and has ∼211 million inhabitants distributed over an area of 8.5 million km². The country is divided into 26 states and the country capital, a federal district (Brazilian Institute of Geography and Statistics - IBGE, n.d.). These 27 states are grouped in 5 administrative macro-regions: North (7 states), Northeast (9 states), Center-West (4 states), Southeast (4 states) and South (3 states). The country is located in a tropical region and has 3 main Köppen climate types and 12 subtypes (Alvares et al., 2013).

### 2.3 Outcome and covariates

Our outcome is a dengue hospitalization event. We used the Brazilian Hospital Admission System (SIH), a nationwide database that comprises individual level data of all hospitalizations covered by the Universal Healthcare System (SUS) in Brazil. We defined a dengue hospitalisation by the following ICD-10 codes: ‘A90’, ‘A91’, ‘A97’, ‘A970’, ‘A971’, ‘A972’, ‘A979’ (Coelho et al., 2016). We build daily time series aggregating the number of events by date of hospitalisation for each municipality of residence.

We used the number of dengue cases as a covariate. We used the National Dengue Surveillance System (SINAN-Dengue), that receives any notification for a suspected or confirmed dengue case. We selected the confirmed cases of dengue and build daily times times series aggregating the number of cases by date of symptoms onset at each municipality. To add this covariate in the model in a sensitivity analysis, we derived the 7-days moving average of the dengue cases series, an estimated time of progression from symptoms to hospitalization.

Both databases are publicly accessible and following ethically agreed principles on open data, the use of this data did not require ethical approval in Brazil, according to the Brazilian Ethics Resolution n° 510/2016.

### 2.4 Temperature exposure assessment

Our exposure of interest is the daily average temperature at each Brazilian municipality. To derive it, we used the hourly 2-metres temperature (gridded 0.1° x 0.1°) from reanalysis products (ERA5-Land), freely available by the Copernicus Climate Service through the Climate date Store (Muñoz-Sabater et al., 2021). We estimated the daily mean temperature for each municipality by calculating the mean daily temperature of each grid cell and weighting it to the municipality area. All these weighted mean areas were done with the ‘exactextractr’ R package (Baston, Daniel, 2023).

Regarding the performance of the temperature estimated from ERA5-Land reanalysis in Brazil, one study evaluated the agreement between ERA5-Land and monitoring stations for monthly temperature averages, using 12 automatic stations for the period 2011-2020 from one Brazilian state in the Northeast region (Pernambuco state - PE), with an average R-square of 0.92 (Araújo et al., 2022). For this study, we performed a validation comparing the daily average of temperature from monitoring stations at the municipal level with the ERA5-Land estimates described before. We used data from 389 stations (269 automatic and 120 manual) from the National Institute of Meteorology (National Institute of Meteorology (INMET), 2024), with at least 90% of days with complete data during the period. The analysis of 340 municipalities covering the 27 states and approximately 65 million individuals showed a Pearson correlation coefficient of 0.94, R-square of 0.90 and RMSE of 1.54°C. Further information is available in the supplementary material.

### 2.5 Data Analysis

We used a two-stage approach in the times-series analysis. At the first stage, we fitted Conditional Poisson Models for each Brazilian state (Armstrong et al., 2014; Gasparrini, 2021). At the second stage, we pooled the 27 estimates using a multivariate meta-analysis (Gasparrini et al., 2012; Gasparrini and Armstrong, 2013; Jackson et al., 2011).

#### 2.5.1 First-stage analysis

At the first-stage, we run the following model for each state:

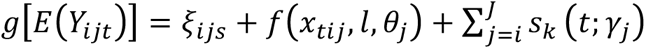

Where *Y_ijt_* is the daily count of cases by the *j-th* state on the *i-th* municipality, ξ_ijs_ the month- municipality strata term conditioned out, f(x_tij_, l, θ_j_) is the bi-dimensional exposure-lag-response distributed lag non-linear cross-basis for the mean temperature on the *j-th* state by each day of delay, until 21 days of lags (Gasparrini et al., 2015; Martínez-Solanas and Basagaña, 2019). The cross-basis is parametrized with natural splines, with 2 knots equally spaced on the dose-response structure and 3 knots equally spaced on the log transformed scale for the lag-response structure. The last term is the long-term trend model choice for temperature trend along the whole period, a natural spline with 7 degrees of freedom for each year over the whole period.

#### 2.5.2. Second-stage analysis

We ran a multivariate meta-analysis with random effects, so from the *j-th* state-level study, θ_j_, we have the following equation:

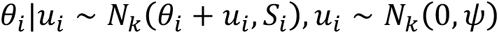

Where *θ* is the study to estimate, with *u*_*i*_ the random effects for the coefficients of this study together with *S*_*j*_ covariance matrix of intra state studies and *ψ* covariance matrix of between states studies. We run one meta-analysis for the whole country and another for each of the macro administrative regions separately. For the whole country meta-analysis, the *θ* studies were taken from all the 27 states, and for the macro regions meta-analyses, *θ* was only taken from states pertaining to the same macro region.

#### 2.5.3 Sensitivity analyses

We ran three sensitivity analyses to explore the DLNM parametrization and to account for the number of dengue cases. On the first sensitivity analysis, we parameterized the cross-basis with 3 knots equally spaced on the dose-response structure and, as in the main analysis, 3 knots equally spaced on the log transformed scale for the lag-response structure. The second sensitivity analysis we parameterized the cross-basis with 2 knots equally spaced on the dose-response structure as the main analysis, however with 4 knots placed on the days 1, 2, 7, 14 for the lag-response structure. On the third sensitivity analysis we adjusted the first stage models adding the 7-days moving average of the times series of confirmed dengue cases, considering the average time between incubation period and time to hospitalization.

#### 2.5.4 Reporting

We report the overall cumulative relative risk (RR) compared to the Minimum Hospitalisation Temperature (MHT) point. MHT is defined as the temperature which has an overall RR equal to 1. We also report RR for the lag effects up to 21 days at the 50th and 95th percentiles of the temperature distribution.

All the analyses were run on R Software, version 4.1.2.

## 3. Results

### 3.1 Climate data description

A descriptive table (**Table S1**) of the mean daily temperature for each state, macro regions and the whole country over the ten years period is shown in the supplementary material. For the whole period of ten years, it varied from −0.09 °C to 34.8 °C. **Figure 1** gives the daily mean temperature distribution across all states.

**Figure 1.**
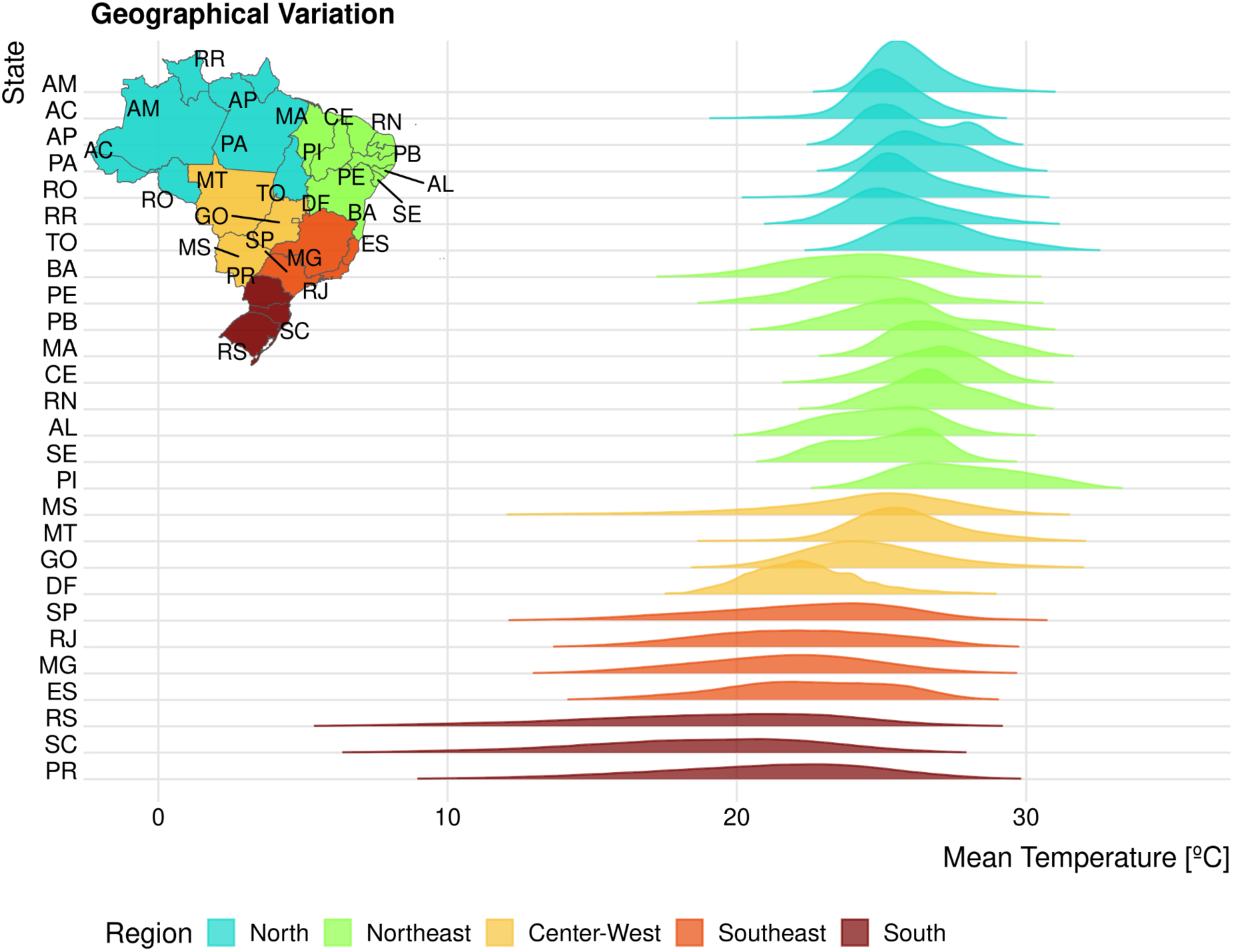
Mean temperature density distribution over each state. On y-axis is each of the state acronyms and on x-axis is the range of mean temperature in degree Celsius. The height of each curve is the cumulative density of mean temperature at that specific degree. The colours are given by each macro administrative region. The inset map gives the location of each of the state code at Brazilian geography.

### 3.2 Dengue hospitalization data description

We excluded dengue hospitalizations from five small municipalities (e.g, small islands), because there was no ambient temperature for them. This filtering resulted in a loss of 153 (0.03%) hospitalized cases, for the whole period of the 10 years. The final time series aggregation by municipalities encompasses a total of 579,703 hospital admissions due to dengue over the 10 years’ period of 2010 to 2019 in 5,565 municipalities.

**Table 1** gives a characterization of the data summarised by each macro administrative region. Overall, the mean age for a hospitalisation by dengue was 30 years old. Between regions it can be seen a difference of 12 years old to the mean age of hospitalisation, being the mean age of hospitalisation of 37 years old for the South region and 25 years old for the Northeast region. The sex ratio is typically representative of the sex ratio estimates from the national statistics. The crude in-hospital mortality in general is low, with mortality rate of 0.6% of the total hospitalizations, and there are some discrepancies between regions. The Southeast has a rate of 1.0% of the hospitalizations with the highest death outcome rate. The North and Northeast with lowest rate of mortality with an average of 0.4% rate of deaths to the total hospitalizations. **Figure S1** gives a visual description of the time series for dengue hospitalisation for each region.

**Table 1.** Characteristics description of Dengue Hospitalisation data.

| Variables | N | Brazil (N=579,703) | North (N=75,304) | Northeast (N=224,085) | Center-West (N=100,056) | Southeast (N=155,405) | South (N=24,853) |
| --- | --- | --- | --- | --- | --- | --- | --- |
| Mean Age (IQR), years | 579,703 | 30 (15, 49) | 27 (15, 44) | 25 (12, 45) | 36 (20, 53) | 34 (17, 54) | 37 (21, 55) |
| Age Categories, n (%) years | 579,703 |  |  |  |  |  |  |
| 0 to 1 |  | 8,752 (1.5%) | 1,309 (1.7%) | 4,457 (2.0%) | 1,058 (1.1%) | 1,726 (1.1%) | 202 (0.8%) |
| 1 to 9 |  | 73,507 (13%) | 9,401 (12%) | 37,354 (17%) | 8,438 (8.4%) | 16,620 (11%) | 1,694 (6.8%) |
| 10 to 17 |  | 91,786 (16%) | 12,562 (17%) | 41,991 (19%) | 12,155 (12%) | 22,171 (14%) | 2,907 (12%) |
| 18 to 39 |  | 192,819 (33%) | 29,153 (39%) | 72,769 (32%) | 34,330 (34%) | 48,009 (31%) | 8,558 (34%) |
| 40 to 59 |  | 126,059 (22%) | 14,863 (20%) | 39,640 (18%) | 26,825 (27%) | 38,268 (25%) | 6,463 (26%) |
| 60 to 79 |  | 71,395 (12%) | 6,737 (8.9%) | 22,314 (10.0%) | 14,656 (15%) | 23,494 (15%) | 4,194 (17%) |
| 80+ |  | 15,385 (2.7%) | 1,279 (1.7%) | 5,560 (2.5%) | 2,594 (2.6%) | 5,117 (3.3%) | 835 (3.4%) |
| Self-Reported Race, n (%) | 396,624 |  |  |  |  |  |  |
| Black |  | 12,467 (3.1%) | 1,042 (2.3%) | 3,946 (2.7%) | 1,187 (1.9%) | 5,760 (4.9%) | 532 (2.6%) |
| Pardo |  | 244,869 (62%) | 39,925 (87%) | 120,476 (81%) | 34,973 (55%) | 45,113 (38%) | 4,382 (21%) |
| Indigenous |  | 1,092 (0.3%) | 306 (0.7%) | 116 (<0.1%) | 595 (0.9%) | 62 (<0.1%) | 13 (<0.1%) |
| White |  | 129,031 (33%) | 3,893 (8.5%) | 19,700 (13%) | 24,396 (39%) | 65,746 (55%) | 15,296 (74%) |
| Asian |  | 9,165 (2.3%) | 700 (1.5%) | 4,003 (2.7%) | 2,116 (3.3%) | 1,991 (1.7%) | 355 (1.7%) |
| (Missing) |  | 183,079 | 29,438 | 75,844 | 36,789 | 36,733 | 4,275 |
| Sex, n (%) | 579,703 |  |  |  |  |  |  |
| Female |  | 310,308 (54%) | 38,225 (51%) | 121,175 (54%) | 54,916 (55%) | 82,550 (53%) | 13,442 (54%) |
| Male |  | 269,395 (46%) | 37,079 (49%) | 102,910 (46%) | 45,140 (45%) | 72,855 (47%) | 11,411 (46%) |
| Duration of Hospitalisation, median (IQR), days | 579,703 | 3 (2, 4) | 2 (2, 3) | 3 (2, 4) | 2 (2, 3) | 3 (2, 4) | 2 (2, 3) |
| In-hospital mortality, n (%) | 579,703 | 3,436 (0.6%) | 281 (0.4%) | 958 (0.4%) | 548 (0.5%) | 1,510 (1.0%) | 139 (0.6%) |

### 3.4 First-stage Results

The cumulative RR over all lags compared to the MHT on each state level, for the whole period of analysis, 2010 to 2019 are given in the supplementary material (**Figure S2**).

### 3.5 Second-stage Results

**Figure 2 and 3**, presents the cumulative RR over all lags compared to the MHT derived from the meta-analysis, for the whole country and for each macro-region. **Figure 2B and 2C**, presents the lag effects for the 50th (23.96° C) and 95th (28.68° C) percentiles of mean temperature distribution for Brazil. The lag effects for each macro-region is shown on supplementary material, **Figure S3, S4, S5, S6** and **S7**. A summary of the RR and MHT for Brazil and each macro-region is shown in **Table 2**.

**Figure 2.**
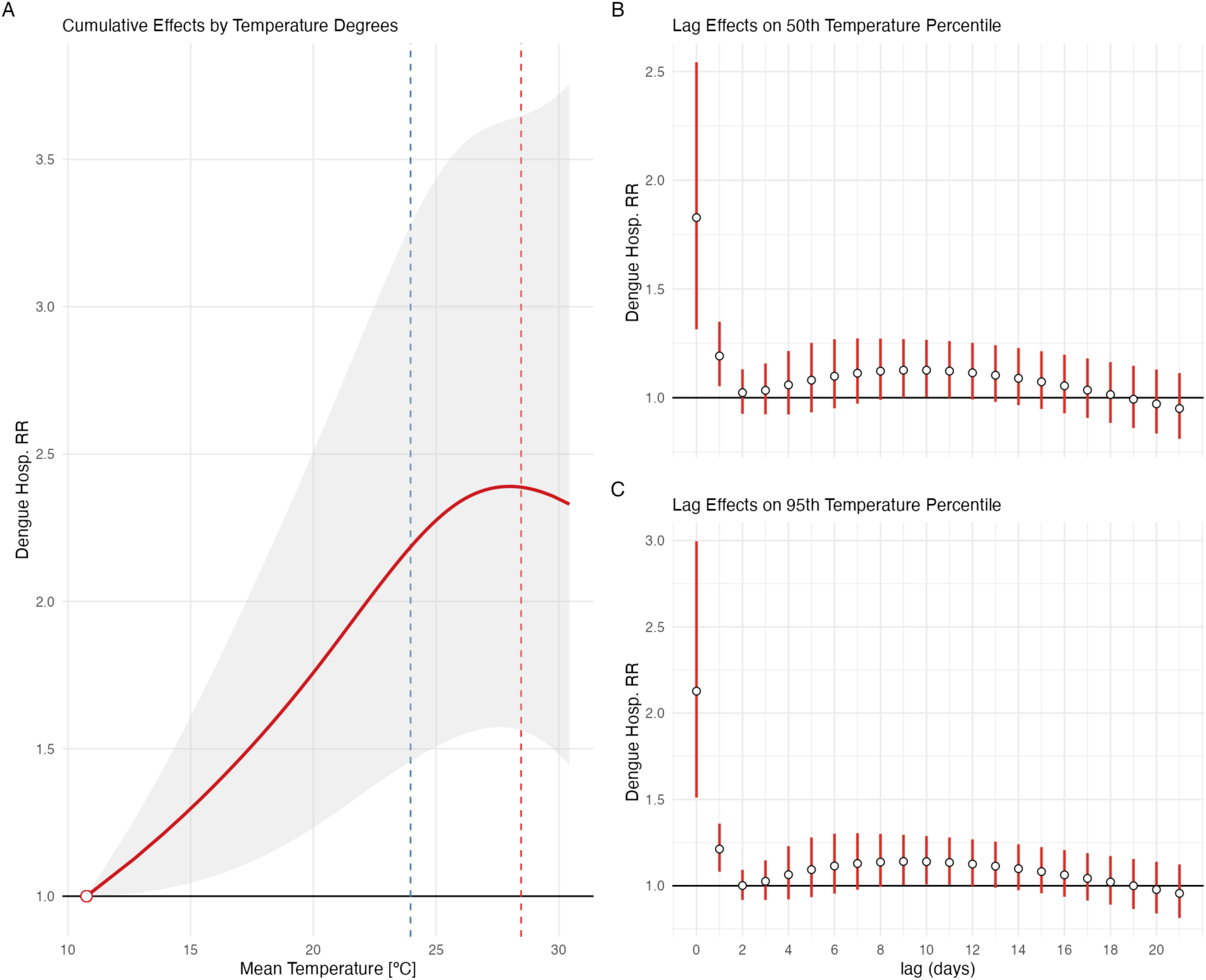
Pooled cumulative relative risk for the whole country meta-analysis. A) Cumulative RR over all lags for a Dengue hospitalisation relative to the MHT. Vertical traced lines mark the 50th (Blue) and 95th (Red) percentile of the temperature distribution. The grey shade ribbon is 95% confidence interval derived from the meta-analysis. B) Lag effect to the RR to the MHT of Hospitalisation due to Dengue at the 50th (23.96° C) percentile of temperature. C) Lag effect to the RR to the MHT of hospitalisation due to Dengue on the 95th (28.68° C) percentile of temperature. MHT: Minimum Hospitalisation Temperature; RR: relative risk.

**Figure 3.**
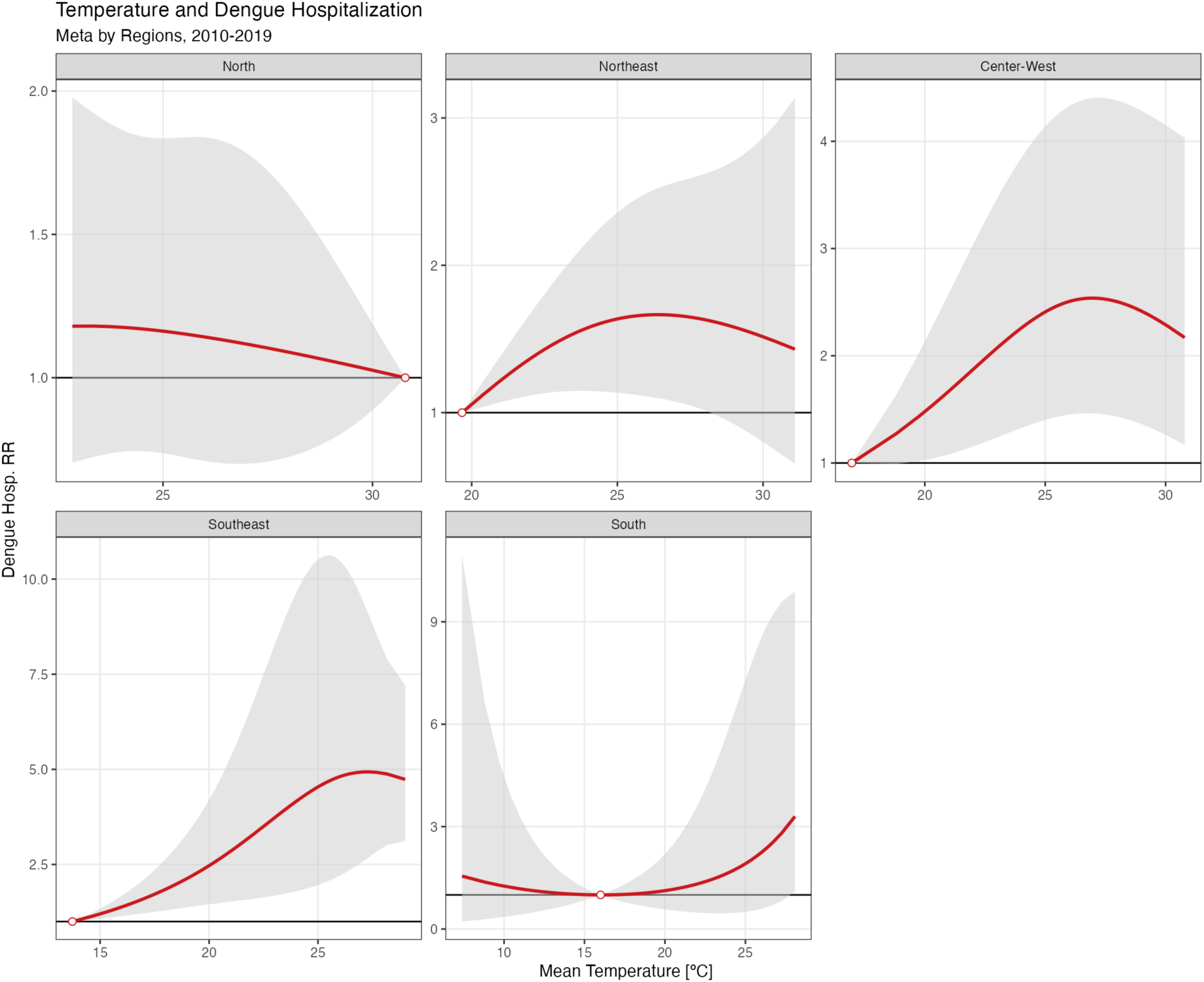
Pooled cumulate relative risk to each of the macro region meta-analysis. Cumulative overall lags RR to the MHT curves to temperature by region. Each panel is given by a meta-analysis model run over all the states coefficients and covariance matrices that pertains to a given macro administrative region. Order by region is given by latitude extent. MHT: Minimum Hospitalisation Temperature; RR: relative risk.

**Table 2.** Dengue hospitalisation relative risk (RR) over Brazil and each macro-region.

| Unit | MHT (°C) | RR (IC 95%) 50th<br>(23·96° C) | RR (IC 95%) 95th<br>(28·68° C) |
| --- | --- | --- | --- |
| Brazil | 10.8 | 2.185 (1.457-3.276) | 2.385 (1.556-3.655) |
| North | 30.8 | 1.139 (0.707-1.836) | 1.048 (0.811-1.355) |
| Northeast | 19.7 | 1.660 (1.116-2.468) | 1.547 (0.860-2.784) |
| Center-West | 17 | 2.393 (1.394-4.110) | 2.420 (1.365-4.289) |
| Southeast | 13.7 | 3.335 (1.605-6.929) | 4.930 (2.567-9.468) |
| South | 16 | 1.119 (0.580-2.159) | 2.253 (0.592-8.569) |

RR: Relative Risk to the Minimum Hospitalisation Temperature of hospitalisation due Dengue
MHT: Minimum Hospitalisation Temperature, on Celsius degree

### 3.6 Sensitivity analyses

From the meta-analyses, on the 50th percentile for whole Brazil the RR was 2.080 (95% CI 1.210-3.578) and 2.315 (95% CI 1.558-3.439), for the first and second sensitivity analyses parametrizations, respectively. After adjusting for the number of dengue cases (third sensitivity analysis), the RR on the 50th percentile for whole Brazil was 1.991 (95% CI 1.382-2.867).

Overall, the sensitivity analyses result for each macro-region were comparable with the main analysis at the 50^th^ percentile (**Figure 4**) and at the 95th percentile (**Figure S8**), except for the South region. There was an increased risk for dengue hospitalization in the South in the three sensitivity analyses, particularly for the 95th percentile, although with high imprecision (RR 4.830, 95% CI, 1.34617.331). Additional results are shown on **Table S2** and **Figures S9**, **S10, S11,** and **S12**, in the supplementary material.

**Figure 4.**
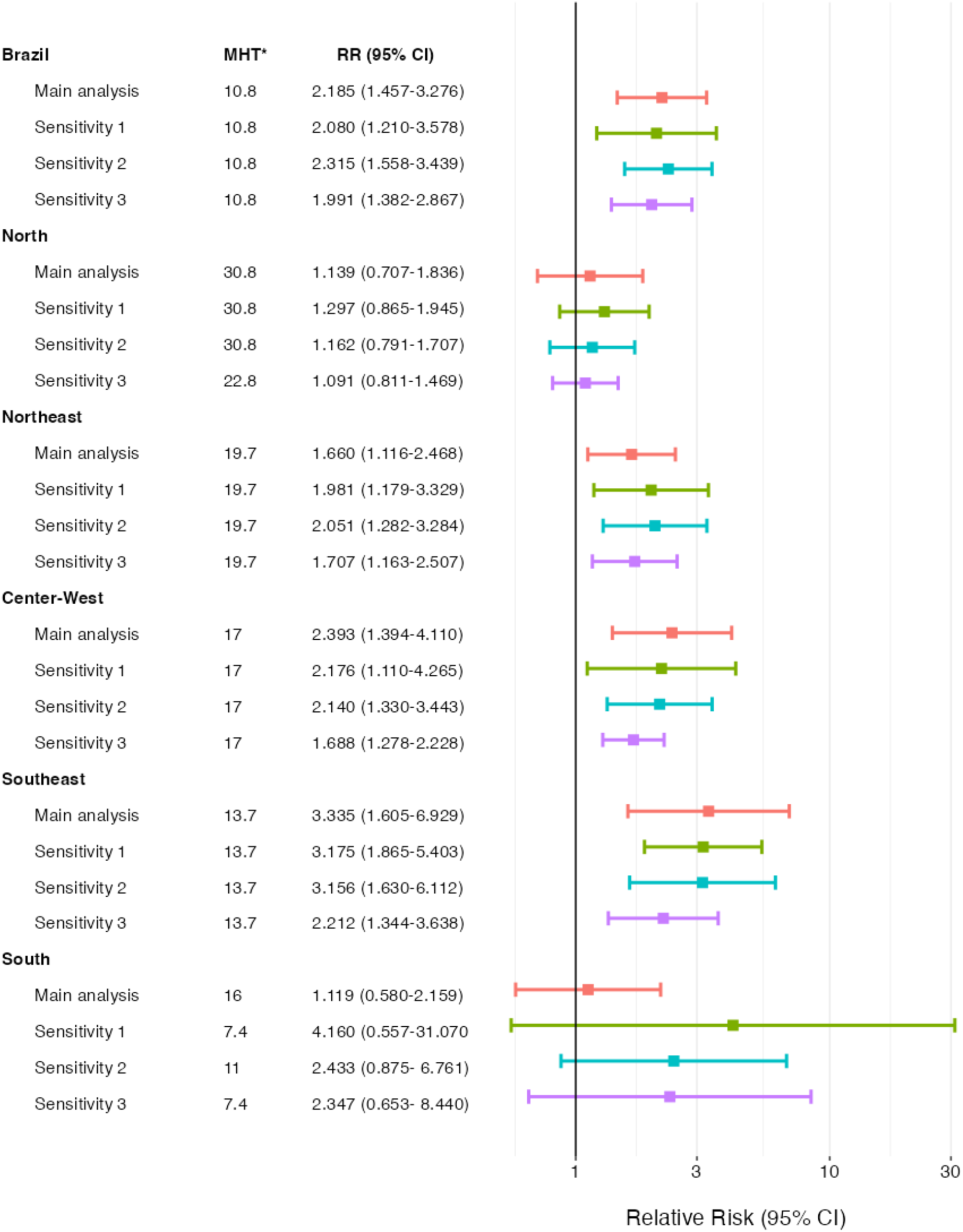
Dengue hospitalization relative risk by Brazil and each macro-region: main and sensitivity analyses at 50th percentile of temperature. *MHT: Minimum Hospitalisation Temperature, on Celsius degree; RR: relative risk

## 4. Discussion

We ran a two-stage time stratified design study to the association between ambient temperature and hospitalisation due to Dengue. Our main finding is an increased risk of being hospitalised by Dengue as the temperature increases. In a quantitative way, in general for Brazil, above 20 degrees Celsius, the RR to the MHT is 1.2x higher. This is the cumulative effects of lags up to 21 days before the date of hospitalisation. We found a stronger immediate effect on the RR to the hospitalisation. We found the same patterns for each macro region of Brazil, but the North and South regions.

In comparison with the literature, we see that, in general, higher temperature is related to an augmented RR of all-cause and cause-specific hospitalisation and mortality (Jacobson et al., 2021; Martínez-Solanas and Basagaña, 2019; Pudpong and Hajat, 2011; Silveira et al., 2019; Wang et al., 2021; Zhao et al., 2019). There is scarce literature on the association between temperatures and hospitalisation by an infectious disease. Our study is the first, to our knowledge, to measure an association between temperature and hospitalisation caused by dengue. Interestingly, we did not observe a typical U-shape curve increase on the risk for hospitalization, as commonly observed for studies looking for all-cause and cause-specific hospitalisations, except for the South region, where the association was unclear. The pattern we observed has been reported for non-infectious causes in the US and China (Vaidyanathan et al., 2019; Wang et al., 2021). Some specific features of dengue could explain this difference. First, its progression to severe disease is highly linked with factors that would be aggravated as the temperature increases, such dehydration and capillary leak syndrome. Second, the incidence of the dengue disease, of any severity, is much lower during cold months, therefore limiting the power to observe a potential effect of cold temperatures on Dengue hospitalizations. This reflects an additional challenge when studying climate variables and vector-borne infectious diseases severity, among others (Imai et al., 2015).

Overall, in the country and each of the macro regions, the extreme heat (above 20 Celsius degrees) has an effect of increasing the severity of cases, which evolves to a necessity of a hospitalisation. The association for the North and South regions are unclear and a direct comparison between regions is not straightforward because of regional disparities as well as climate disparities. Although Brazil has a universal access health system, SUS has great disparities over geographical distribution (Noronha et al., 2020). Thus, we might expect that regions with more hospital beds per population, such as South and Southeast, would hospitalise milder cases compared with regions with fewer hospital beds. This is also important during massive dengue outbreaks, where the threshold for hospitalisation might change according to beds availability and age distribution of cases.

Several mechanisms can play a role in the observed overall association and lag effects. The immediate effect generating the first peak on lag-0 is expected and observed in the majority of studies evaluating temperature and heat waves and hospitalizations and deaths (Morral-Puigmal et al., 2018; Pudpong and Hajat, 2011; Royé et al., 2020; Saha et al., 2014; Schwartz et al., 2004; Vaidyanathan et al., 2019). It reflects an immediate worsening of the clinical condition due to high temperature over the course of an infection. This sharp increase on lag-0 reinforces that our results is a direct effect of ambient temperature on hospitalisation risk, rather than the indirect effect of temperature on mosquito activity which increases risk of dengue infection, association usually present in the scale of months (Lowe et al., 2021). Even though, the sensitivity analysis when adding the 7-day rolling mean of mild cases as a covariate in the model shows that the effect of temperature on our model is reduced from 2.185 to 1.991 at the 50^th^ and from 2.385 to 2.221 at the 95^th^ percentile of temperature (**Table S2**). This sensitivity analysis suggests, as before, that the augmented risk for hospitalization by dengue with increased temperature it is a direct effect of heat on individuals already infected by dengue. This finding shows that high temperature seems have a short- and a long-term effect on dengue burden, being the first particularly on severity and the second on infection.

We hypothesize that the natural evolution of 1 to 3 days of incubation period from an infection, plus 5 to 7 days to the complete clearance of the viral infection, can be affected by heat exposure based on the lagged effects observed in our results. If an individual is exposed to higher temperatures during the viral infection evolution this can lead to more severe infection, as seen on lagged-effect around 5 to 7 days before hospitalization. Additionally, the RR decrease right after the lag0-1 effect could be explained by depletion of susceptibles (“harvesting effect”) (Saha et al., 2014; Schwartz et al., 2004). This mechanism is reported for the lag between temperature and deaths, but also has been hypothesised in another study evaluating temperature and other seasonal infectious diseases such as hand, foot and mouth disease (Yi et al., 2020). Importantly, the lag-effect varies over regions, for instance, the South region has a greater delayed lag-effect than other regions. However, when varying the knots placement in a sensitivity analysis, lag-effects were less prominent, increasing the likelihood of collinearity explaining this finding (Basagaña and Barrera-Gómez, 2022).

Our study has some strengths. We evaluated a nationwide database providing a 10-year time-series of dengue hospitalizations in a LMIC exposed to a wide temperature range. We used ambient temperature from reanalysis products from ERA5-Land (Royé et al., 2020), which might increase the generalizability of our results to many other LMIC countries as countries in the regions which dengue has been expanding during last years. Finally, we used the DLNM approach, accounting for dose-response and lag-response structures and correlated daily data on temperature (De Schrijver et al., 2022; Gasparrini, 2021).

Our study has limitations to be mentioned. We did not evaluate other factors that could modify the high temperature effects, such as green space, urbanisation and relative humidity. In the same extension, we did not evaluate individual factors, such as age and sex, that could show different effects of temperature and risk of dengue hospitalization in vulnerable populations.

Second, we obtained estimates for 27 states and pooled them for Brazil and each corresponding macro-regions. States from the same macro-region will have similar climate as well as a similar dengue incidence. This choice of aggregation allows to comparing states with probably similar transmission conditions to the mosquito as well hospitalizations conditions (Lee et al., 2021).

However, this may introduce bias to the analysis and do not allow for generalizations to some municipalities within each state or to border line areas. Finally, we might have exposure misclassification by using the ERA-5-land reanalysis, particularly for the North region based on our validation. Nevertheless, the use of daily average temperature from ERA5-Land reanalysis has been applied in several epidemiological analysis (Alahmad et al., 2023; Kephart et al., 2022), with two studies showing minimal differences in estimates when comparing the exposure from ERA5-Land reanalysis with the observed weather station data for mortality (Mistry et al., 2022; Royé et al., 2020). One of these studies included 18 municipalities from Brazil in the period 1997-2011, with comparable estimates of excess mortality due to cold (2.83, 95% CI, 2.29-3.38 for station versus 2.90, 95% CI, 2.07-3.69 for ERA5-Land) and to heat (0.73, 95% CI, 0.47-0.99 for station versus 0.70, 95% CI, −0.14-1.50 for ERA5-Land) (Mistry et al., 2022). Thus, we did not expect meaningful impact on the results due to the potential exposure misclassification.

## 5. Conclusion

In conclusion, we observed an association between high temperature and increased risk of dengue hospitalization in Brazil. That association is mainly driven by an immediate effect of heat, and varied over the five Brazilian macro-regions, being unclear at the North and South regions. This study adds to the gap in knowledge showing a short-term effect of temperature on dengue severity, in addition to the well-known association between long-term temperature and dengue infection reported in the literature.

## Acknowledgements

The authors also thank the research funding agencies: the Coordenação de Aperfeiçoamento de Pessoal de Nível Superior -- Brazil (Finance Code 001 to RLPS), Conselho Nacional de Desenvolvimento Científico e Tecnológico -- Brazil (grant number: 141698/2018-7 to RLPS). OTR is funded by a Sara Borrell fellowship (CD19/00110) from the Instituto de Salud Carlos III. We acknowledge support from the grant CEX2018-000806-S funded by MCIN/AEI/ 10.13039/501100011033, and support from the Generalitat de Catalunya through the CERCA Program. The funding agencies had no role in the conceptualization of the study.

## Data Availability Statement

All data used in this study are publicly available. Hospitalisation data is available at http://sihd.datasus.gov.br/principal/index.php, and temperature at https://cds.climate.copernicus.eu/#!/home. The code to reproduce this analysis is available at: https://github.com/rafalopespx/dengue_t2m_severity_paper

## Author Contribution Statement

**Rafael Lopes:** Conceptualization, Data curation, Formal analysis, Funding acquisition, Methodology, Software, Visualization, Writing – original draft

**Xavier Basagaña:** Methodology, Supervision, Validation, Writing - review & editing

**Leonardo S. L. Bastos:** Data curation, Writing - review & editing

**Fernando A. Bozza:** Writing - review & editing

**Otavio T. Ranzani:** Conceptualization, Data curation, Formal analysis, Funding acquisition, Methodology, Supervision, Validation, Writing - review & editing

## Conflicts of interest

None.

## Supplementary Material

### ERA5-Land validation

We performed a validation comparing the daily average of temperature from monitoring stations at the municipal level with the ERA5-Land estimates. We used data from 389 stations (269 automatic and 120 manual) from the National Institute of Meteorology (INMET), with at least 90% of days with complete data during the period. This validation has a total of 1,221,051 days evaluated, from 340 municipalities covering the 27 states and the 5 macro-regions of Brazil.

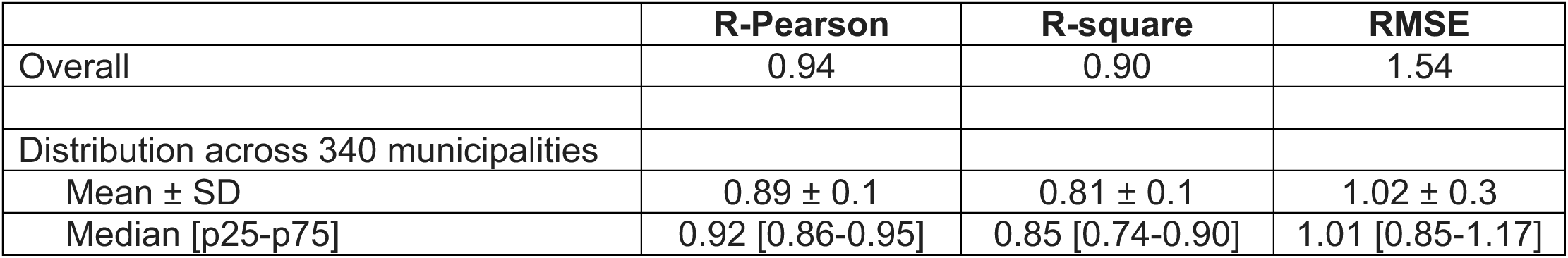

|  | **R-Pearson** | **R-square** | **RMSE** |
| --- | --- | --- | --- |
| Overall | 0.94 | 0.90 | 1.54 |
| Distribution across 340 municipalities |  |  |  |
| Mean ± SD | 0.89 ± 0.1 | 0.81 ± 0.1 | 1.02 ± 0.3 |
| Median [p25-p75] | 0.92 [0.86-0.95] | 0.85 [0.74-0.90] | 1.01 [0.85-1.17] |

Spatial distribution of R-square for each of the 340 municipalities. The shade areas are the municipality areas. N stands for North (n=39), NE for Northeast (n=95), CW for Center-West (n=42), SE for Southeast (n=103) and S for South (n=61).

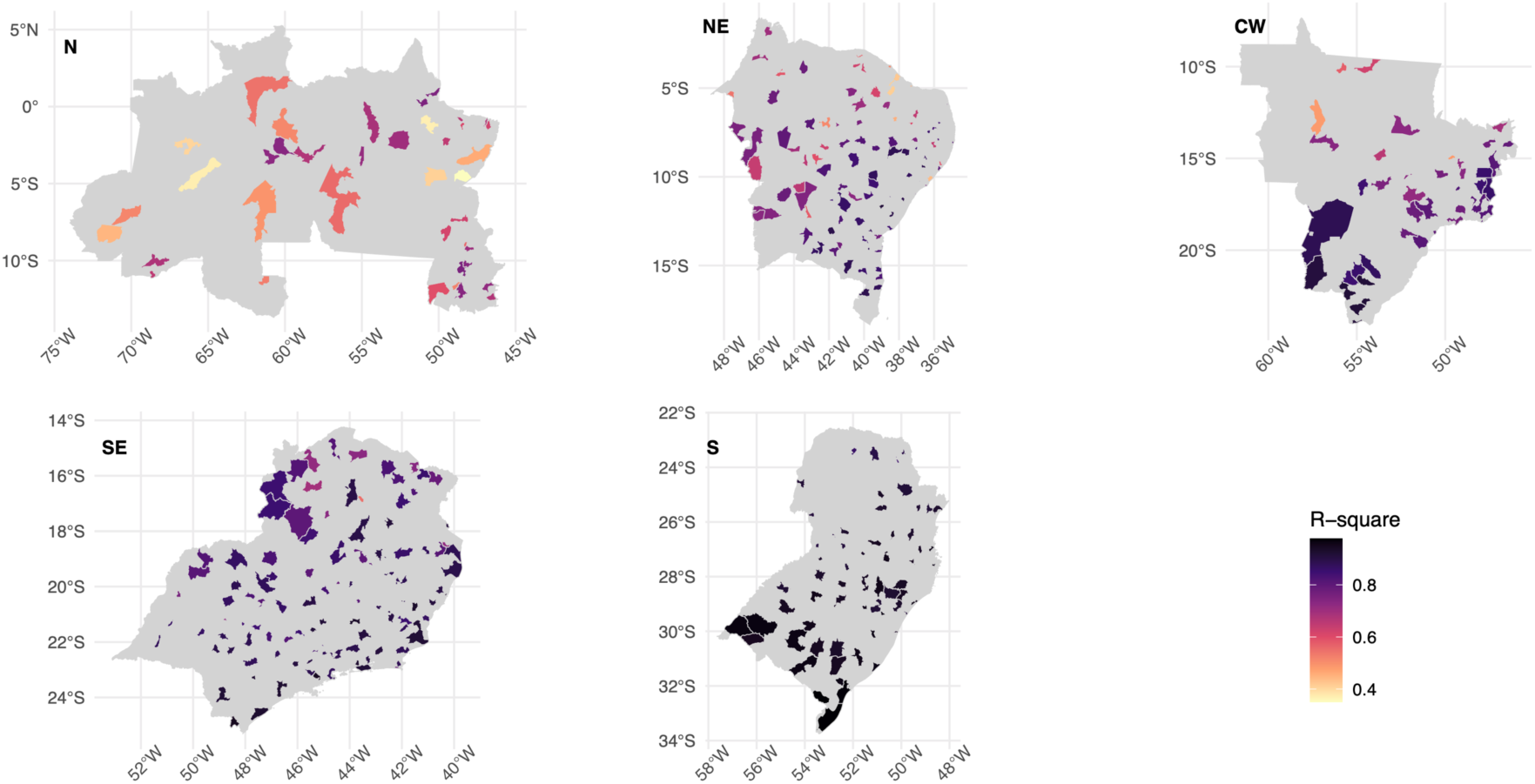

Spatial distribution of RMSE for each of the 340 municipalities. The shade areas are the municipality areas. N stands for North (n=39), NE for Northeast (n=95), CW for Center-West (n=42), SE for Southeast (n=103) and S for South (n=61).

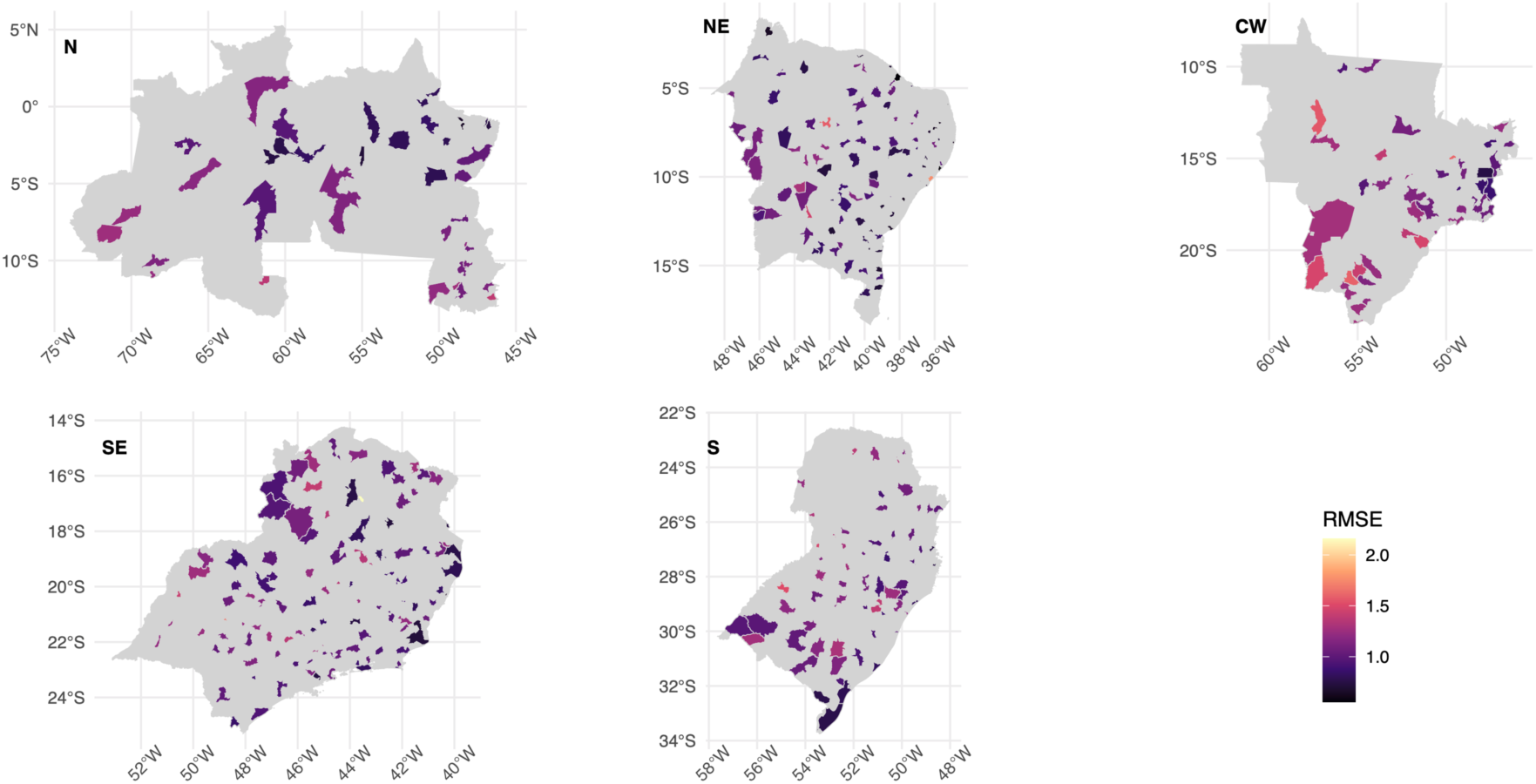

**Table S1.** Descriptive distribution of 2m height Mean Temperature.

| Brazil, Region, State | Minimum | 5th | 25th | 50th | 75th | 95th | Maximum |
| --- | --- | --- | --- | --- | --- | --- | --- |
| Brazil | -0.09 | 15.39 | 20.92 | 23.96 | 26.08 | 28.68 | 34.8 |
| North | 13.99 | 23.91 | 25.15 | 26.13 | 27.39 | 29.32 | 34.32 |
| AC | 13.99 | 22.81 | 24.35 | 25.13 | 26.01 | 27.49 | 31.01 |
| AM | 17.98 | 24.18 | 25.09 | 25.81 | 26.68 | 28.39 | 33.06 |
| AP | 21.98 | 23.72 | 24.73 | 25.65 | 27.14 | 28.59 | 30.5 |
| PA | 21.7 | 24.24 | 25.38 | 26.31 | 27.53 | 28.99 | 32.59 |
| RO | 14.11 | 23.34 | 24.67 | 25.52 | 26.6 | 28.59 | 31.61 |
| RR | 20.04 | 22.88 | 24.29 | 25.33 | 26.67 | 28.82 | 32.24 |
| TO | 19.6 | 24.17 | 25.58 | 26.71 | 28.03 | 30.27 | 34.32 |
| Northeast | 14.04 | 21.49 | 24.16 | 25.8 | 27.24 | 29.54 | 34.39 |
| AL | 18.6 | 21.76 | 23.52 | 25.02 | 26.32 | 27.9 | 32.78 |
| BA | 14.04 | 19.99 | 22.31 | 24.08 | 25.73 | 27.8 | 33.99 |
| CE | 19.09 | 23.82 | 25.58 | 26.77 | 27.81 | 29.16 | 32.41 |
| MA | 21.75 | 24.53 | 25.79 | 26.84 | 28.09 | 29.89 | 33.29 |
| PB | 19.17 | 22.36 | 24.19 | 25.49 | 26.74 | 29.15 | 32.08 |
| PE | 17.59 | 20.77 | 22.95 | 24.35 | 25.76 | 27.84 | 32.71 |
| PI | 20.71 | 24.44 | 26.07 | 27.55 | 29.3 | 31.41 | 34.39 |
| RN | 20.18 | 24.02 | 25.56 | 26.6 | 27.66 | 29.29 | 31.86 |
| SE | 19.77 | 22.17 | 23.74 | 25.35 | 26.53 | 27.77 | 32.25 |
| Center-West | 6.09 | 20.68 | 23.34 | 24.88 | 26.41 | 28.96 | 34.3 |
| DF | 15.96 | 19.42 | 21.01 | 22.23 | 23.54 | 25.87 | 29.58 |
| GO | 11.3 | 20.99 | 22.97 | 24.4 | 26 | 28.76 | 34.18 |
| MS | 6.09 | 16.95 | 22.42 | 24.71 | 26.46 | 28.81 | 34.26 |
| MT | 10.6 | 22.55 | 24.46 | 25.61 | 26.89 | 29.31 | 34.3 |
| Southeast | 4.69 | 16.13 | 19.7 | 22.13 | 24.31 | 27.03 | 34.8 |
| ES | 10.78 | 17.22 | 20.39 | 22.49 | 24.66 | 26.82 | 31.46 |
| MG | 6.71 | 16.15 | 19.47 | 21.76 | 23.82 | 26.62 | 34.54 |
| SP | 4.69 | 15.88 | 19.96 | 22.66 | 24.83 | 27.45 | 34.8 |
| RJ | 9.78 | 16.75 | 19.79 | 22.12 | 24.45 | 27.2 | 32.25 |
| South | -0.09 | 11.03 | 16.52 | 19.92 | 22.76 | 26.02 | 33.03 |
| PR | 0.67 | 13.25 | 18.19 | 21.27 | 23.78 | 26.78 | 32.42 |
| RS | 0.77 | 9.99 | 15.71 | 19.33 | 22.35 | 25.7 | 33.03 |
| SC | -0.09 | 10.8 | 15.86 | 18.98 | 21.73 | 24.89 | 31.13 |

**Figure S1.**
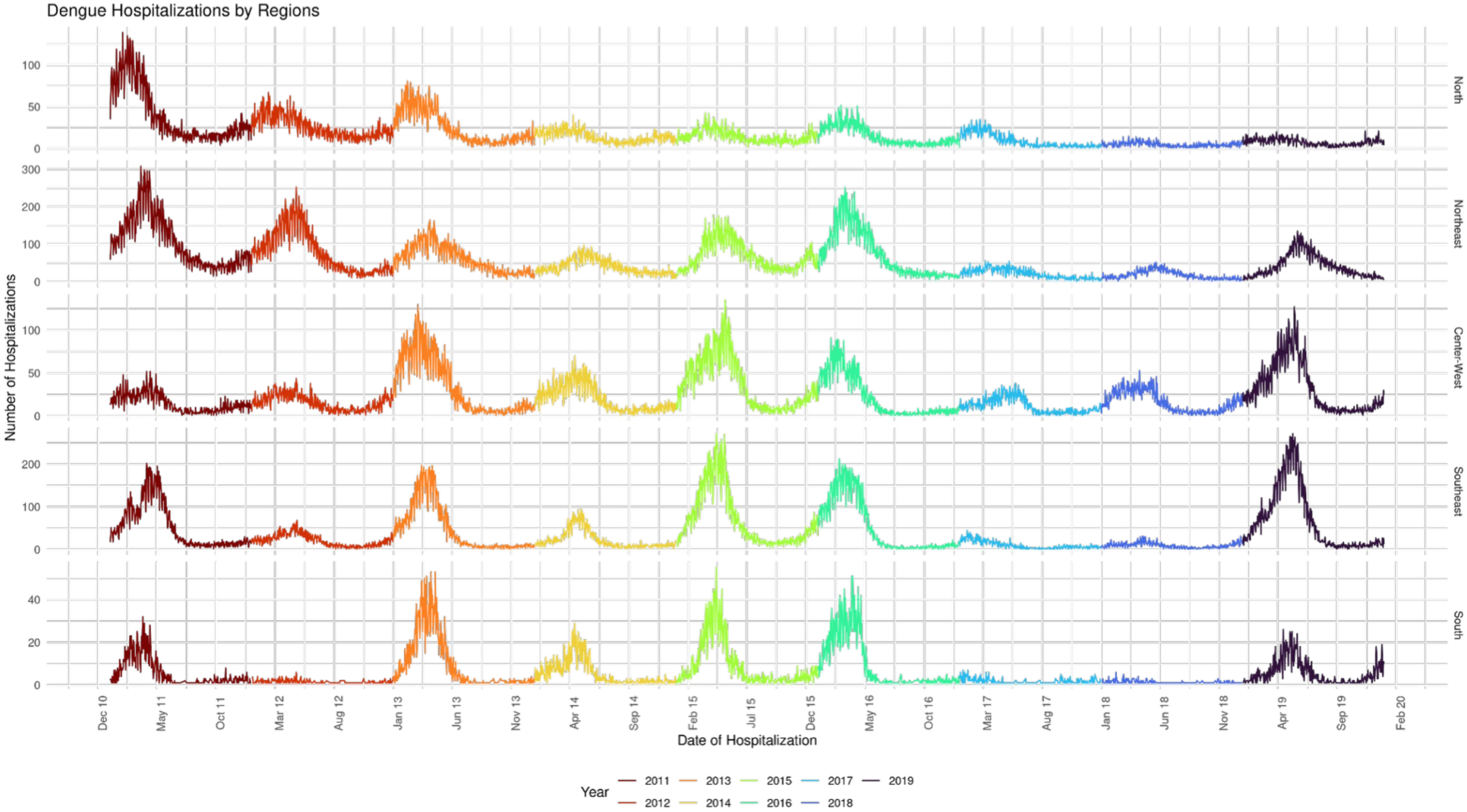
Time series of Dengue hospitalisation by macro administrative region of Brazil. Colour by year, the data covers a period of 10 years, from the whole epidemiological year of 2010 to the whole epidemiological year of 2019.

**Figure S2.**
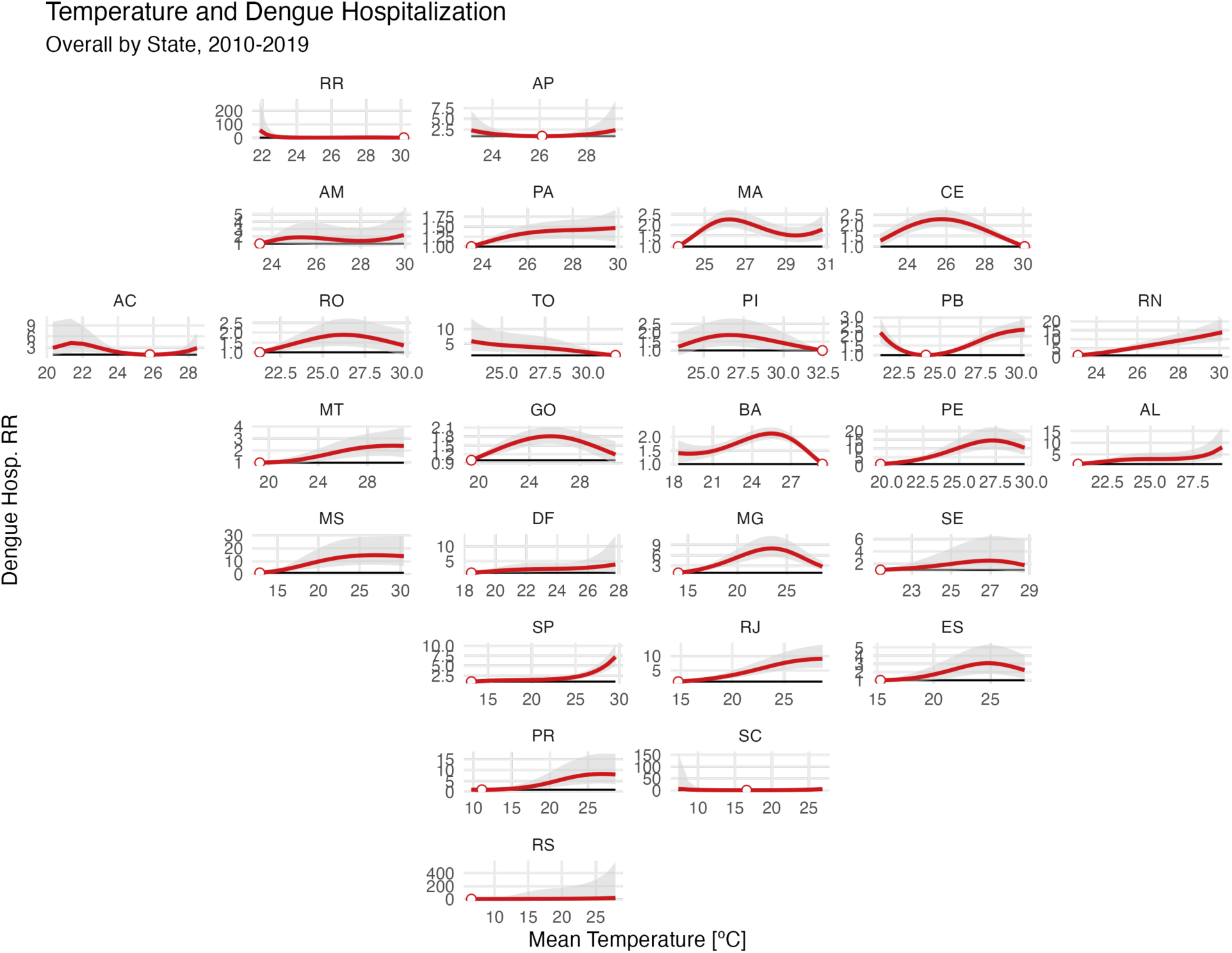
Cumulative relative risk over all the lags compared to the MHT on each state level, for the whole period of analysis, 2010 to 2019 (First stage, Main Analysis)

The curve is plotted in red lines and the 95% confidence interval generated from the fitted model is given by the grey shaded ribbon around it. The title of each subplot is the abbreviations for the name of each state.

**Figure S3.**
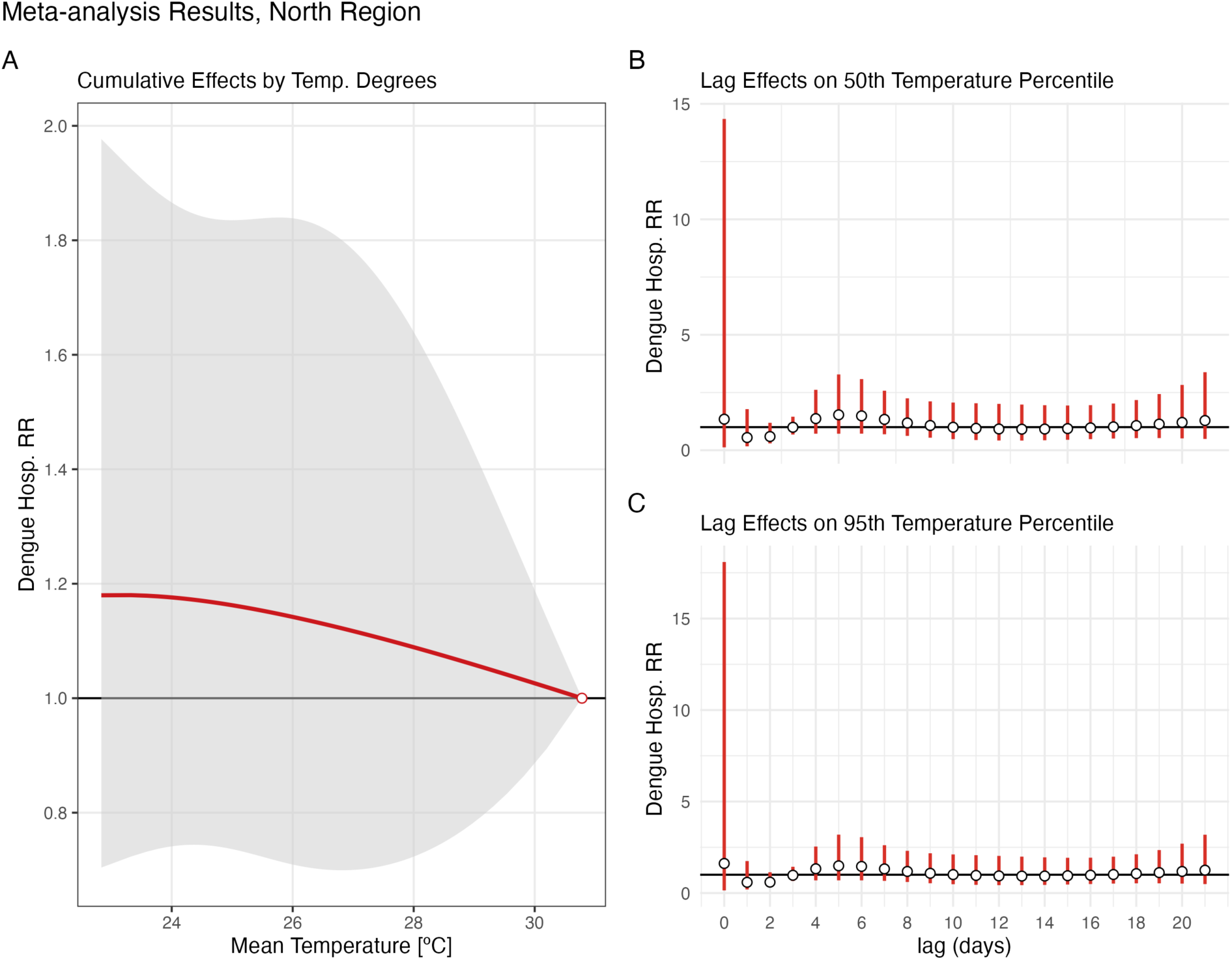
A) Cumulative relative risk over all lags for a Dengue hospitalisation compared to the MHT at the North Region. B) Lag effect of the RR on the 50th percentile of temperature. C) Lag effect of the RR on the 95th percentile of temperature (Main analysis)

The grey shade (A) is 95% confidence interval, as the error bars (B and C), derived from the meta-analysis.

**Figure S4.**
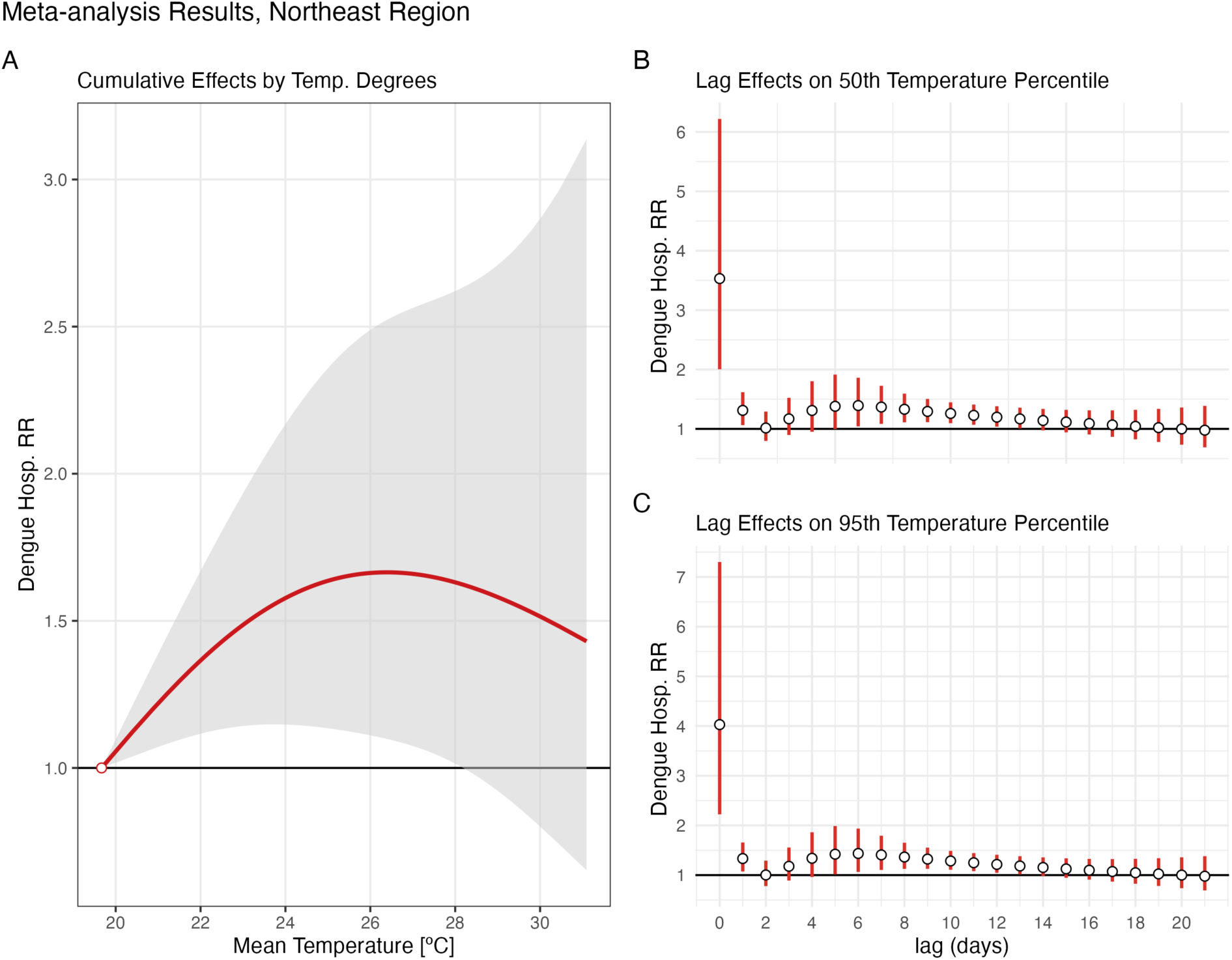
A) Cumulative relative risk over all lags for a Dengue hospitalisation compared to the MHT at the Northeast Region. B) Lag effect of the RR on the 50th percentile of temperature. C) Lag effect of the RR on the 95th percentile of temperature (Main analysis)

The grey shade (A) is 95% confidence interval, as the error bars (B and C), derived from the meta-analysis.

**Figure S5.**
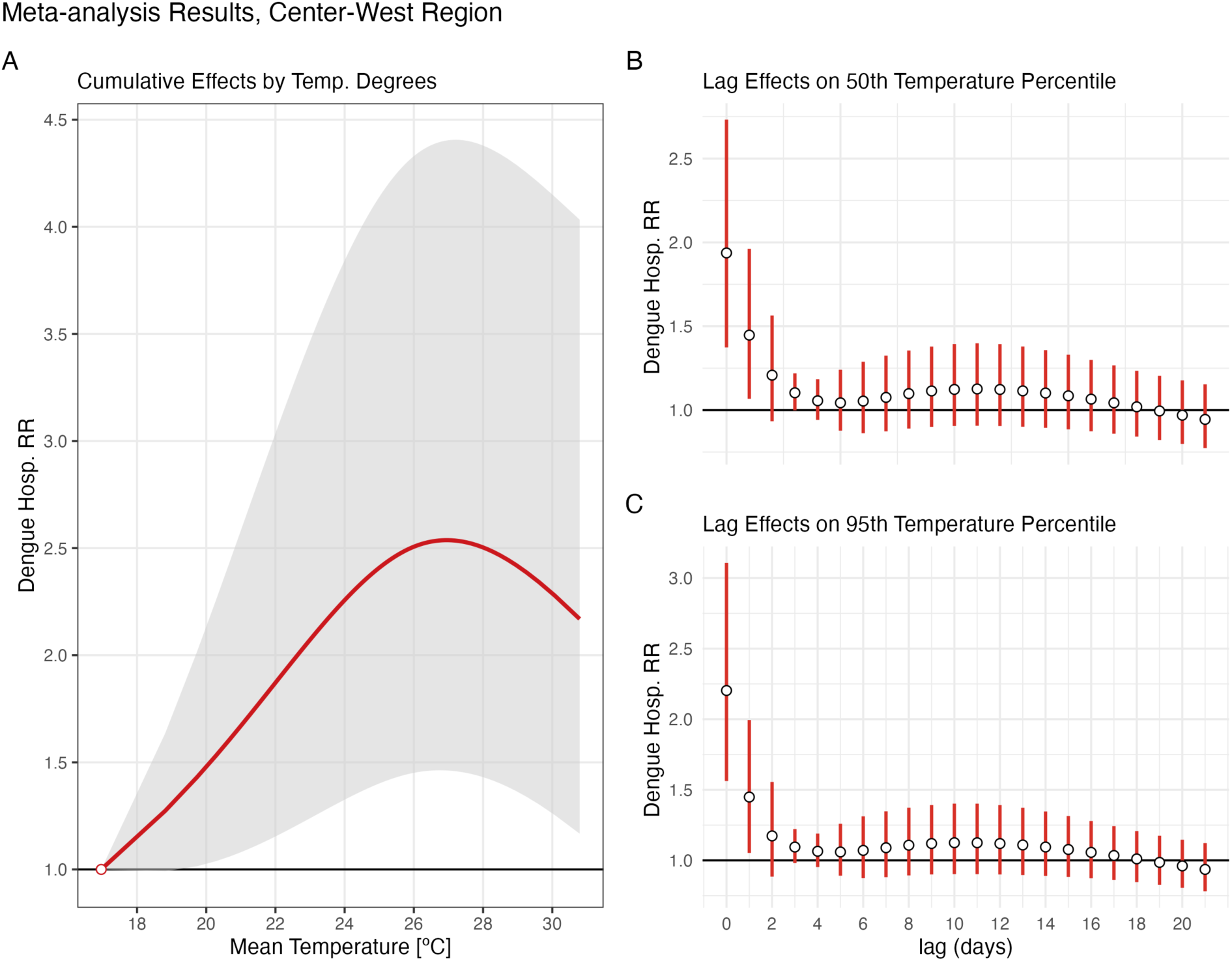
A) Cumulative relative risk over all lags for a Dengue hospitalisation compared to the MHT at the Center-West Region. B) Lag effect of the RR on the 50th percentile of temperature. C) Lag effect of the RR on the 95th percentile of temperature (Main analysis)

The grey shade (A) is 95% confidence interval, as the error bars (B and C), derived from the meta-analysis.

**Figure S6.**
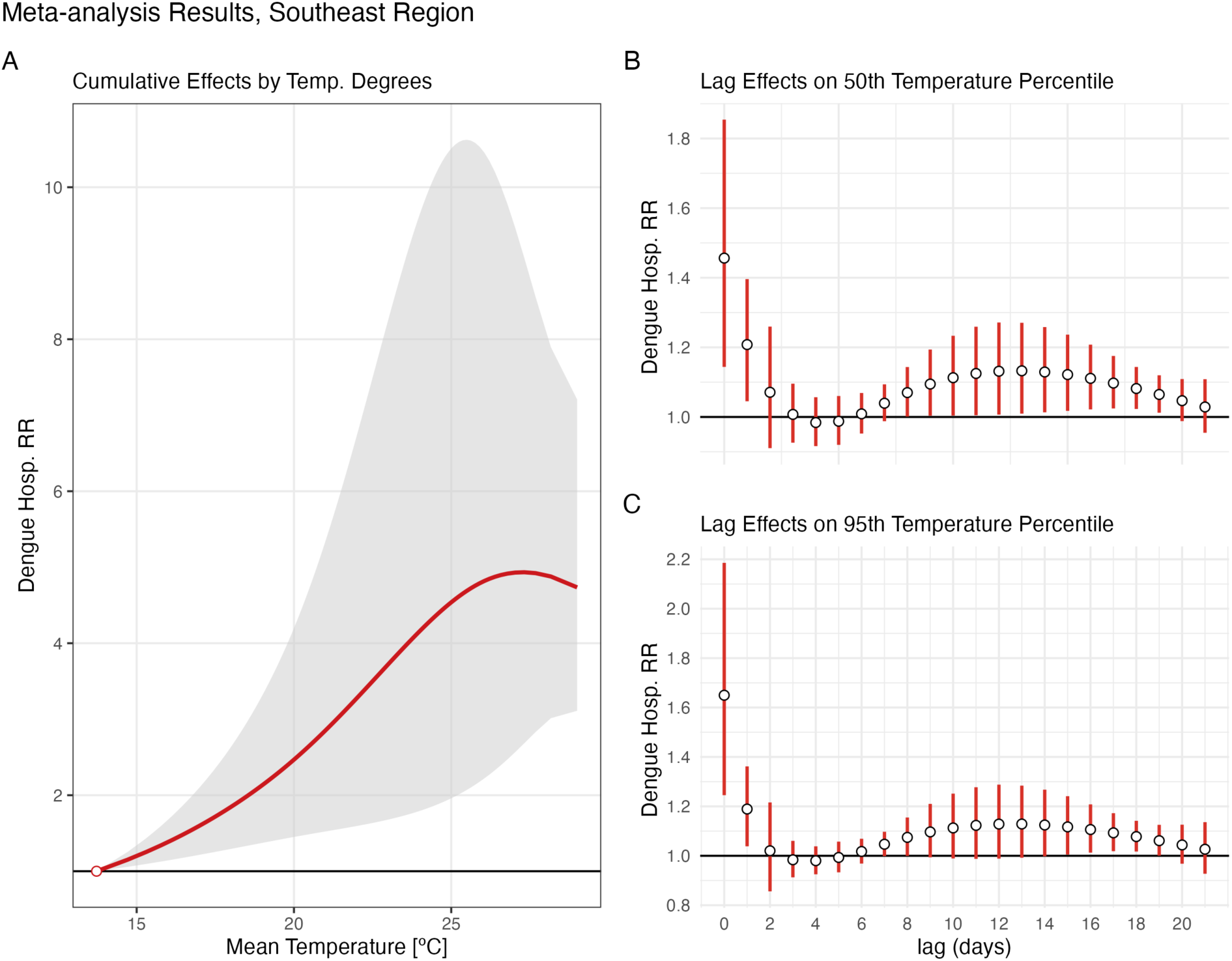
A) Cumulative relative risk over all lags for a Dengue hospitalisation compared to the MHT at the Southeast Region. B) Lag effect of the RR on the 50th percentile of temperature. C) Lag effect of the RR on the 95th percentile of temperature (Main analysis)

The grey shade (A) is 95% confidence interval, as the error bars (B and C), derived from the meta-analysis.

**Figure S7.**
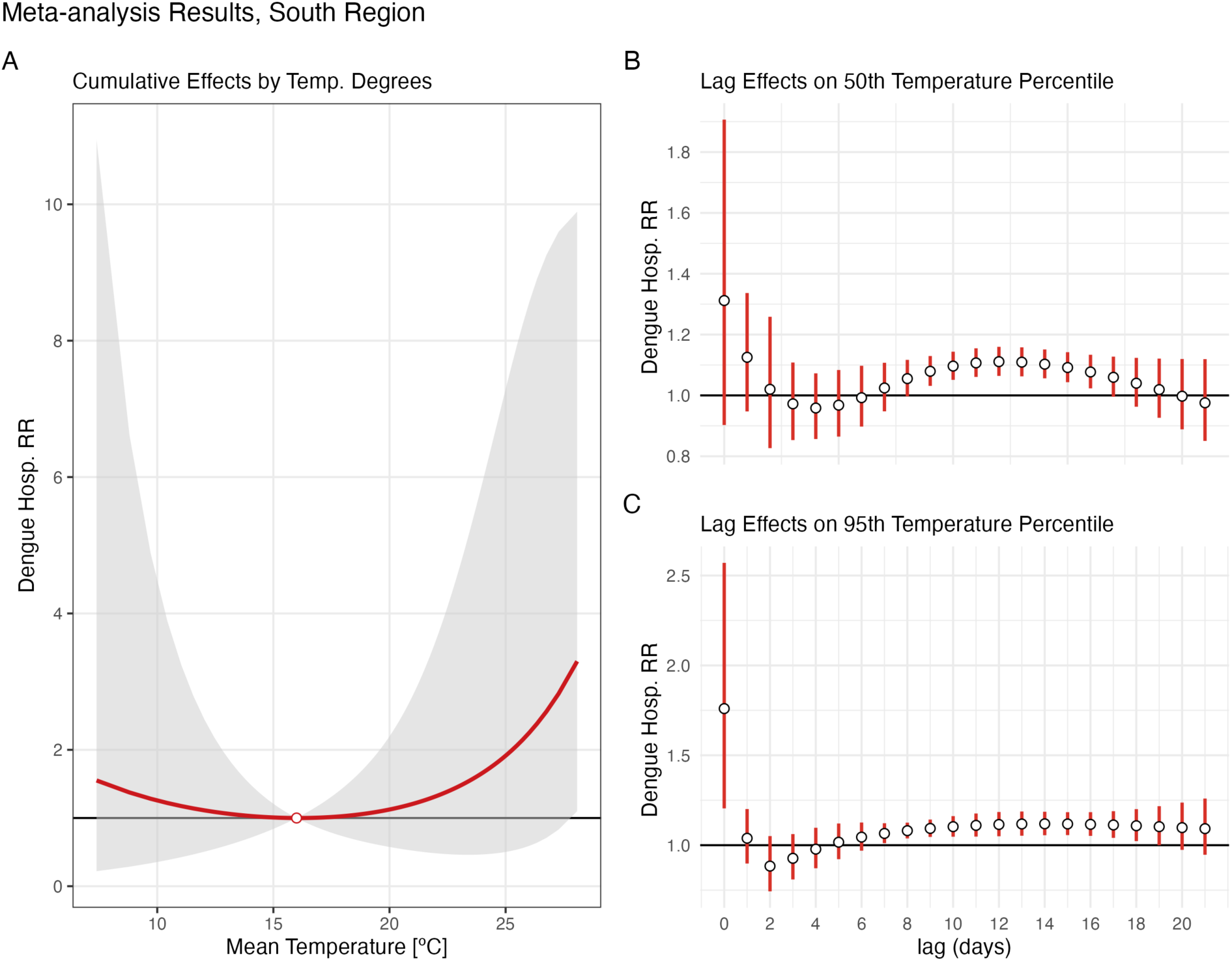
A) Cumulative relative risk over all lags for a Dengue hospitalisation compared to the MHT at the South Region. B) Lag effect of the RR on the 50th percentile of temperature. C) Lag effect of the RR on the 95th percentile of temperature (Main analysis)

The grey shade (A) is 95% confidence interval, as the error bars (B and C), derived from the meta-analysis.

**Table S2.** Dengue hospitalisation relative risk by Brazil and each macro-region: main and sensitivity analyses.

|  | MHT (°C) | RR (95% CI), 50th | RR (95% CI), 95th |
| --- | --- | --- | --- |
| **Main analysis** |  |  |  |
| Brazil | 10.8 | 2.185 (1.457-3.276) | 2.385 (1.556-3.655) |
| North | 30.8 | 1.139 (0.707-1.836) | 1.048 (0.811-1.355) |
| Northeast | 19.7 | 1.660 (1.116-2.468) | 1.547 (0.860-2.784) |
| Center-West | 17 | 2.393 (1.394-4.110) | 2.420 (1.365-4.289) |
| Southeast | 13.7 | 3.335 (1.605-6.929) | 4.930 (2.567-9.468) |
| South | 16 | 1.119 (0.580-2.159) | 2.253 (0.592-8.569) |
| **Sensitivity analysis 1** |  |  |  |
| Brazil | 10.8 | 2.080 (1.210-3.578) | 2.285 (1.152-4.531) |
| North | 30.8 | 1.297 (0.865-1.945) | 1.061 (0.798-1.412) |
| Northeast | 19.7 | 1.981 (1.179-3.329) | 1.983 (0.978-4.020) |
| Center-West | 17 | 2.176 (1.110-4.265) | 2.122 (1.058-4.255) |
| Southeast | 13.7 | 3.175 (1.865-5.403) | 4.358 (2.699-7.038) |
| South | 7.4 | 4.160 (0.557-31.070) | 9.125 (1.308-63.655) |
| **Sensitivity analysis 2** |  |  |  |
| Brazil | 10.8 | 2.315 (1.558-3.439) | 2.501 (1.639-3.819) |
| North | 30.8 | 1.162 (0.791-1.707) | 1.058 (0.824-1.359) |
| Northeast | 19.7 | 2.051 (1.282-3.284) | 2.151 (1.180-3.921) |
| Center-West | 17 | 2.140 (1.330-3.443) | 1.972 (1.172-3.317) |
| Southeast | 13.7 | 3.156 (1.630-6.112) | 3.813 (2.601-5.588) |
| South | 11 | 2.433 (0.875- 6.761) | 5.140 (2.135-12.371) |
| **Sensitivity analysis 3** |  |  |  |
| Brazil | 10.8 | 1.991 (1.382-2.867) | 2.221 (1.512-3.263) |
| North | 22.8 | 1.091 (0.811-1.469) | 1.098 (0.736-1.640) |
| Northeast | 19.7 | 1.707 (1.163-2.507) | 1.735 (1.074-2.803) |
| Center-West | 17 | 1.688 (1.278-2.228) | 1.734 (1.145-2.626) |
| Southeast | 13.7 | 2.212 (1.344-3.638) | 2.553 (1.634-3.989) |
| South | 7.4 | 2.347 (0.653- 8.440) | 4.830 (1.346-17.331) |

RR: Relative Risk to the Minimum Hospitalisation Temperature of hospitalisation due Dengue MHT: Minimum Hospitalisation Temperature, on Celsius degree

**Figure S8.**
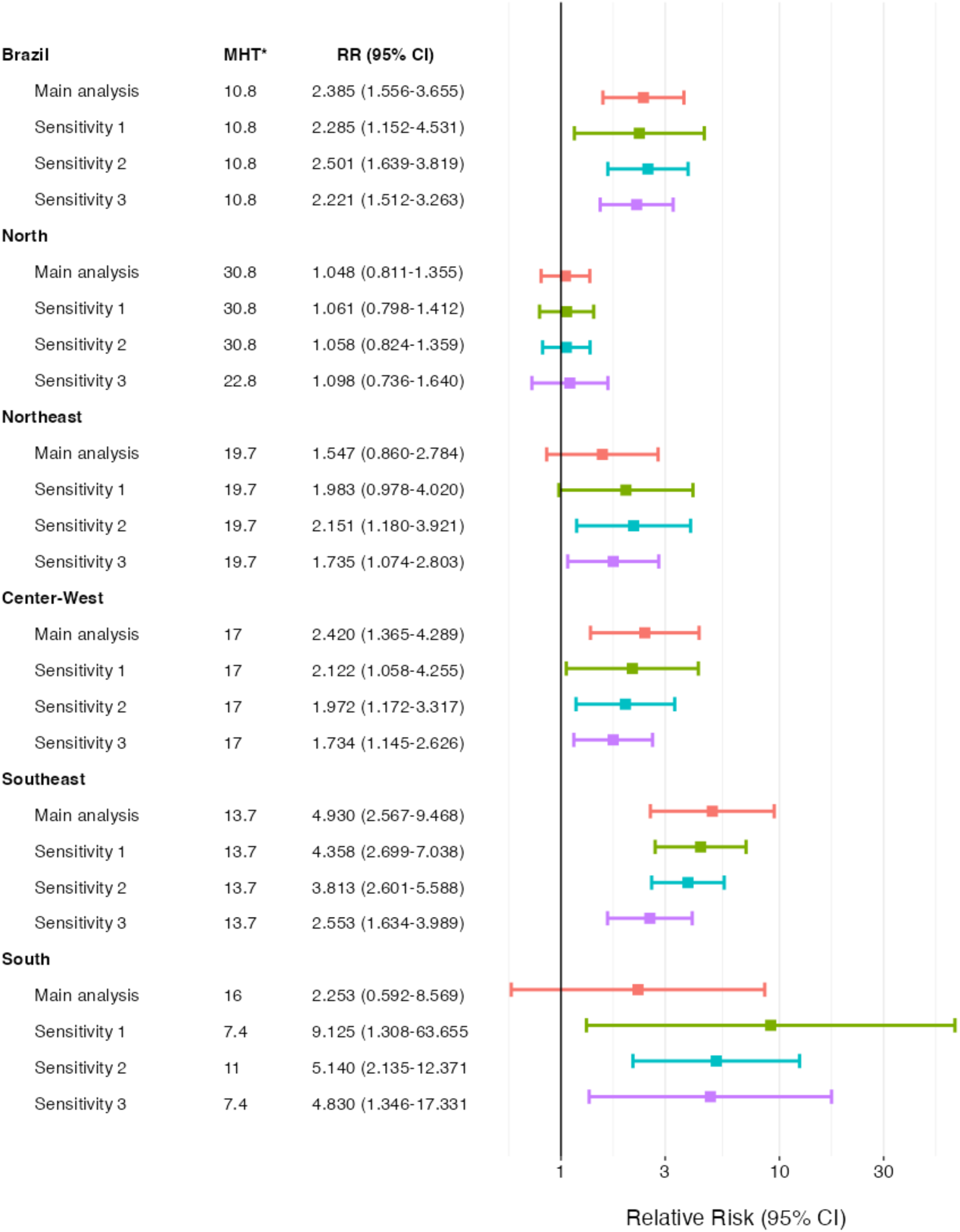
Dengue hospitalisation relative risk by Brazil and each macro-region: main and sensitivity analyses forest plot at 95^th^ percentile of temperature. *MHT: Minimum Hospitalisation Temperature, on Celsius degree

**Figure S9.**
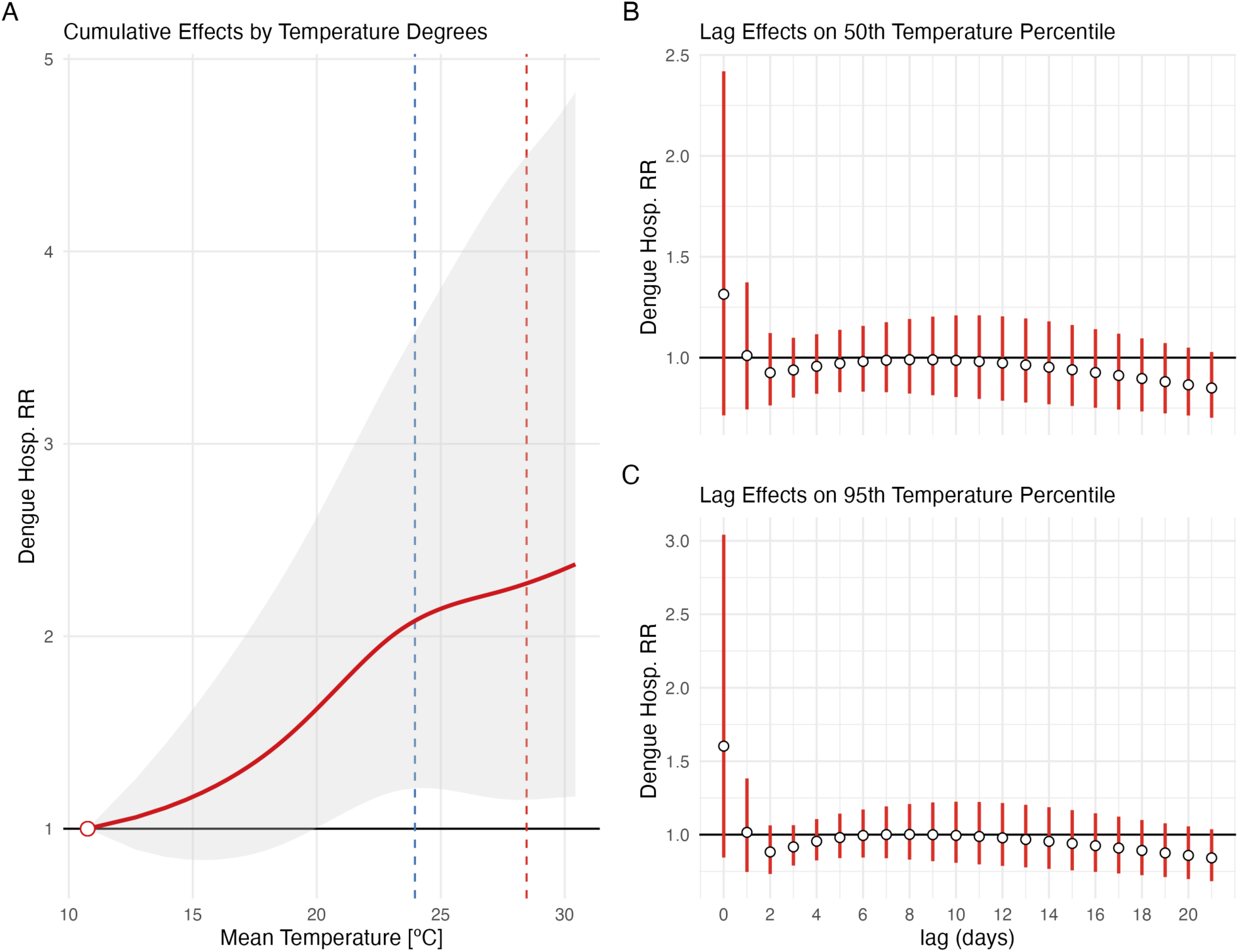
A) Cumulative relative risk over all lags for a Dengue hospitalisation compared to the MHT in Brazil. B) Lag effect of the RR on the 50th percentile of temperature. C) Lag effect of the RR on the 95th percentile of temperature (Sensitivity Analysis 1)

Sensitivity Analysis 1: parametrization of dose-response: 3 knots equally spaced; lag-response: 3 knots equally spaced at the log-scale. Vertical traced lines mark the 50th (Blue) and 95th (Red) percentile of the temperature distribution. The grey shade (A) is 95% confidence interval, as the error bars (B and C), derived from the meta-analysis.

**Figure S10.**
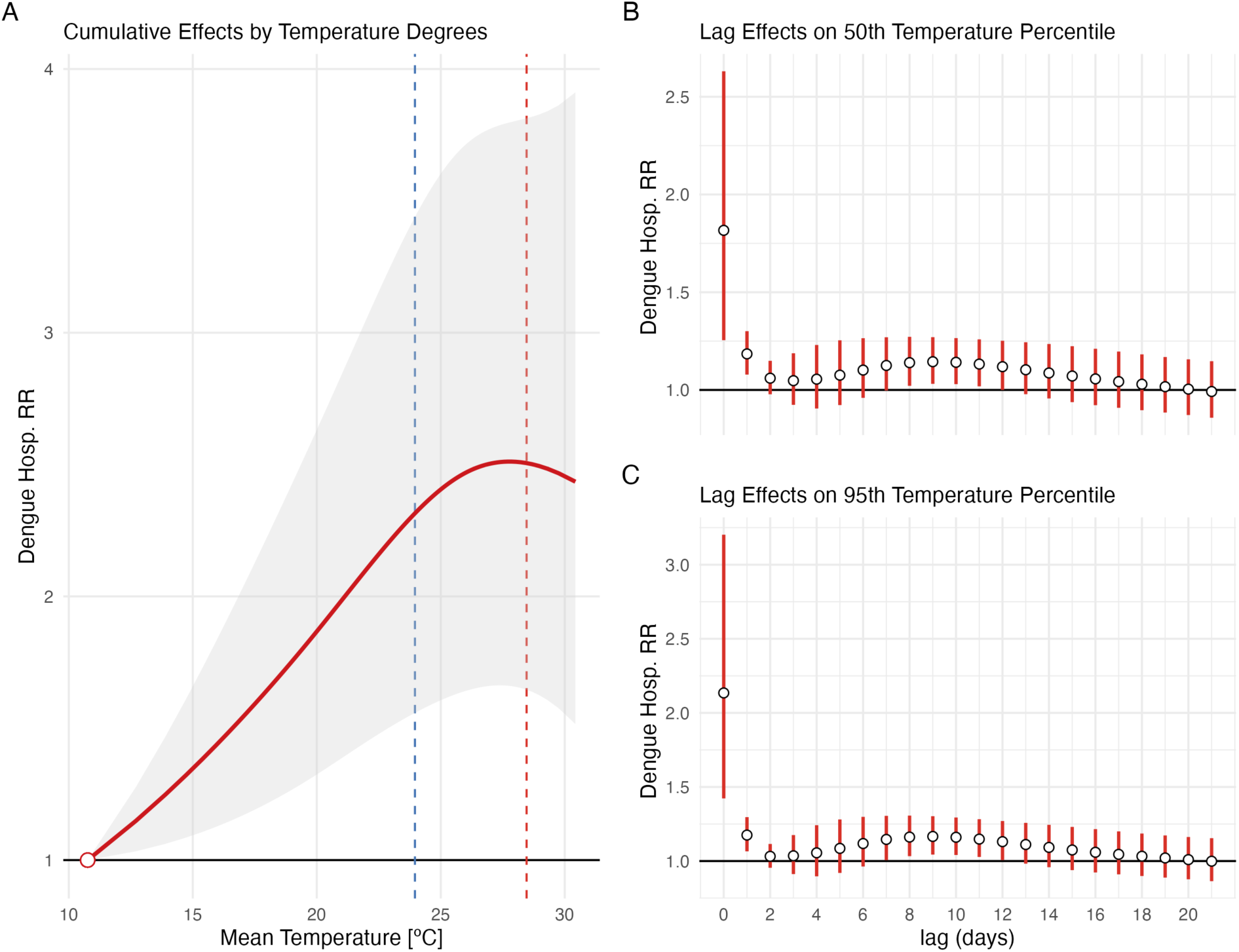
A) Cumulative relative risk over all lags for a Dengue hospitalisation compared to the MHT in Brazil. B) Lag effect of the RR on the 50th percentile of temperature. C) Lag effect of the RR on the 95th percentile of temperature (Sensitivity Analysis 2)

Sensitivity Analysis 2: parametrization of dose-response: 2 knots equally spaced; lag-response: 4 knots at days 1, 2, 7 and 14 from date of hospitalization. Vertical traced lines mark the 50th (Blue) and 95th (Red) percentile of the temperature distribution. The grey shade (A) is 95% confidence interval, as the error bars (B and C), derived from the meta-analysis.

**Figure S11.**
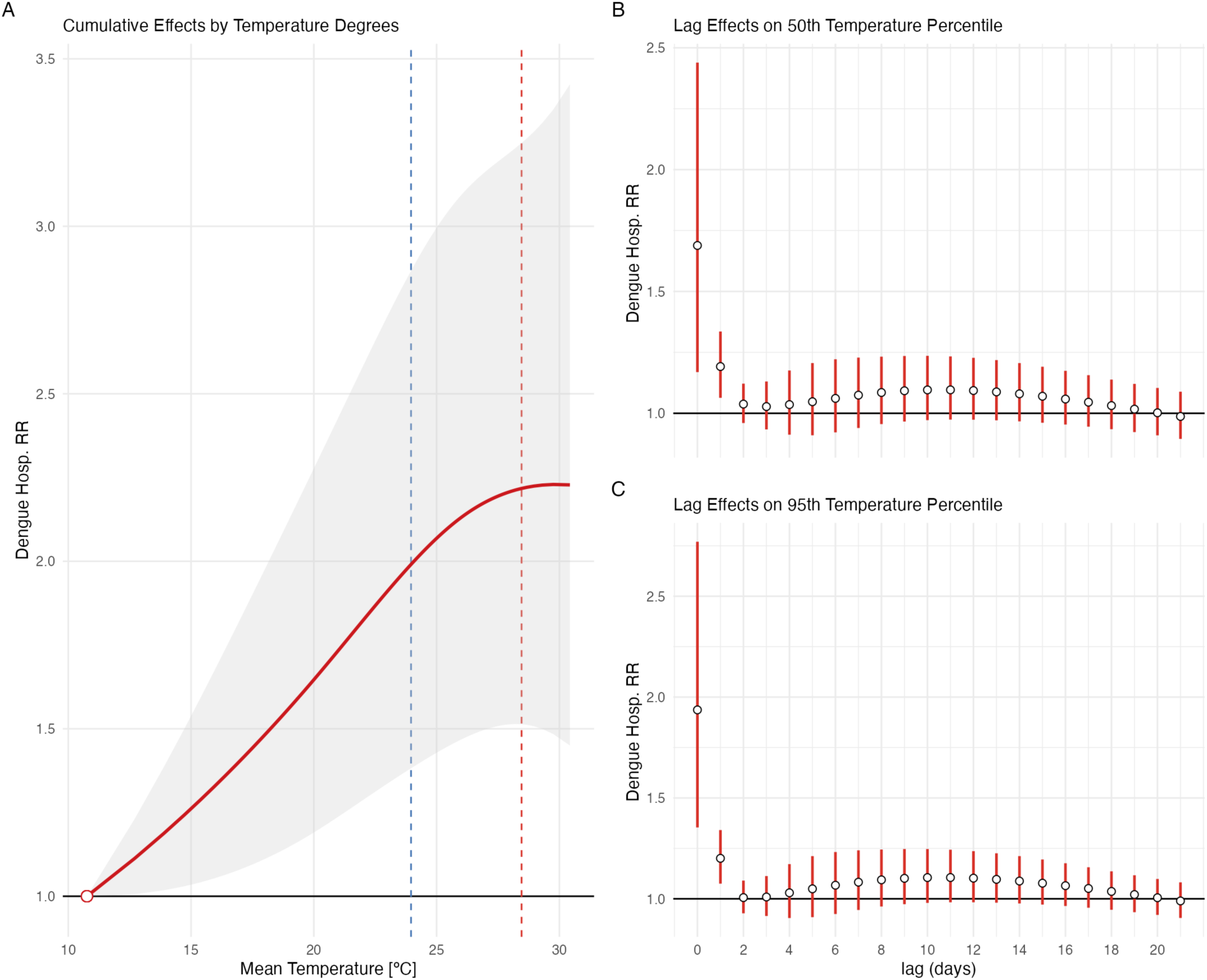
A) Cumulative relative risk over all lags for a Dengue hospitalisation compared to the MHT in Brazil. B) Lag effect of the RR on the 50th percentile of temperature. C) Lag effect of the RR on the 95th percentile of temperature (Sensitivity Analysis 3)

Sensitivity Analysis 3: parametrization of dose-response: 2 knots equally spaced; lag-response: lag-response: 3 knots equally spaced at the log-scale; covariate: 7-day moving average of confirmed dengue cases. Vertical traced lines mark the 50th (Blue) and 95th (Red) percentile of the temperature distribution. The grey shade (A) is 95% confidence interval, as the error bars (B and C), derived from the meta-analysis.

## Notes

### Competing Interest Statement

The authors have declared no competing interest.

### Funding Statement

Conselho Nacional de Pesquisa, Coordenacao Nacional de Aperfeicoamento de Pessoal, Institut de Salud Carlos III.

### Author Declarations

All the data were got from the Brazillian Hospital Admission System (SIH), that can be accessed thourgh the link: - http://tabnet.datasus.gov.br/cgi/deftohtm.exe?sih/cnv/qibr.def

### Summary of Updates

The version of the manuscript has been revised in its structure. New figures were added as well as new numbering of all figures and tables. The manuscript now has page numbers and line numbering.

## References

Alahmad, B., Khraishah, H., Royé, D., Vicedo-Cabrera, A.M., Guo, Y., Papatheodorou, S.I., Achilleos, S., Acquaotta, F., Armstrong, B., Bell, M.L., Pan, S.-C., De Sousa Zanotti Stagliorio Coelho, M., Colistro, V., Dang, T.N., Van Dung, D., De’ Donato, F.K., Entezari, A., Guo, Y.-L.L., Hashizume, M., Honda, Y., Indermitte, E., Íñiguez, C., Jaakkola, J.J.K., Kim, H., Lavigne, E., Lee, W., Li, S., Madureira, J., Mayvaneh, F., Orru, H., Overcenco, A., Ragettli, M.S., Ryti, N.R.I., Saldiva, P.H.N., Scovronick, N., Seposo, X., Sera, F., Silva, S.P., Stafoggia, M., Tobias, A., Garshick, E., Bernstein, A.S., Zanobetti, A., Schwartz, J., Gasparrini, A., Koutrakis, P., 2023. Associations Between Extreme Temperatures and Cardiovascular Cause-Specific Mortality: Results From 27 Countries. Circulation 147, 35–46. 10.1161/CIRCULATIONAHA.122.061832

Alvares, C.A., Stape, J.L., Sentelhas, P.C., De Moraes Gonçalves, J.L., Sparovek, G., 2013. Köppen’s climate classification map for Brazil. metz 22, 711–728. 10.1127/0941-2948/2013/0507

Araújo, C.S.P.D., Silva, I.A.C.E., Ippolito, M., Almeida, C.D.G.C.D., 2022. Evaluation of air temperature estimated by ERA5-Land reanalysis using surface data in Pernambuco, Brazil. Environ Monit Assess 194, 381. 10.1007/s10661-022-10047-2

Armstrong, B.G., Gasparrini, A., Tobias, A., 2014. Conditional Poisson models: a flexible alternative to conditional logistic case cross-over analysis. BMC Med Res Methodol 14, 122. 10.1186/1471-2288-14-122

Basagaña, X., Barrera-Gómez, J., 2022. Reflection on modern methods: visualizing the effects of collinearity in distributed lag models. International Journal of Epidemiology 51, 334–344. 10.1093/ije/dyab179

Baston, Daniel, 2023. exactextractr: Fast Extraction from Raster Datasets using Polygons. R package.

Bhatt, S., Gething, P.W., Brady, O.J., Messina, J.P., Farlow, A.W., Moyes, C.L., Drake, J.M., Brownstein, J.S., Hoen, A.G., Sankoh, O., Myers, M.F., George, D.B., Jaenisch, T., Wint, G.R.W., Simmons, C.P., Scott, T.W., Farrar, J.J., Hay, S.I., 2013. The global distribution and burden of dengue. Nature 496, 504–507. 10.1038/nature12060

Brazilian Institute of Geography and Statistics - IBGE, n.d. 2010 Population Census. [WWW Document]. URL https://www.ibge.gov.br/en/statistics/social/income-expenditure-and-consumption/18391-2010-population-census.html?edicao=19720&t=sobre (accessed 4.27.21).

Burattini, M.N., Lopez, L.F., Coutinho, F.A.B., Siqueira, J.B., Homsani, S., Sarti, E., Massad, E., 2016. Age and regional differences in clinical presentation and risk of hospitalization for dengue in Brazil, 2000-2014. Clinics 71, 455–463. 10.6061/clinics/2016(08)08

Campbell, K.M., Haldeman, K., Lehnig, C., Munayco, C.V., Halsey, E.S., Laguna-Torres, V.A., Yagui, M., Morrison, A.C., Lin, C.-D., Scott, T.W., 2015. Weather Regulates Location, Timing, and Intensity of Dengue Virus Transmission between Humans and Mosquitoes. PLoS Negl Trop Dis 9, e0003957. 10.1371/journal.pntd.0003957

Coelho, G.E., Leal, P.L., Cerroni, M.D.P., Simplicio, A.C.R., Siqueira, J.B., 2016. Sensitivity of the Dengue Surveillance System in Brazil for Detecting Hospitalized Cases. PLoS Negl Trop Dis 10, e0004705. 10.1371/journal.pntd.0004705

Colón-González, F.J., Sewe, M.O., Tompkins, A.M., Sjödin, H., Casallas, A., Rocklöv, J., Caminade, C., Lowe, R., 2021. Projecting the risk of mosquito-borne diseases in a warmer and more populated world: a multi-model, multi-scenario intercomparison modelling study. The Lancet Planetary Health 5, e404–e414. 10.1016/S2542-5196(21)00132-7

Damtew, Y.T., Tong, M., Varghese, B.M., Anikeeva, O., Hansen, A., Dear, K., Zhang, Y., Morgan, G., Driscoll, T., Capon, T., Bi, P., 2023. Effects of high temperatures and heatwaves on dengue fever: a systematic review and meta-analysis. eBioMedicine 91, 104582. 10.1016/j.ebiom.2023.104582

De Schrijver, E., Bundo, M., Ragettli, M.S., Sera, F., Gasparrini, A., Franco, O.H., Vicedo-Cabrera, A.M., 2022. Nationwide Analysis of the Heat- and Cold-Related Mortality Trends in Switzerland between 1969 and 2017: The Role of Population Aging. Environ Health Perspect 130, 037001. 10.1289/EHP9835

Gasparrini, A., 2021. The Case Time Series Design. Epidemiology 32, 829–837. 10.1097/EDE.0000000000001410

Gasparrini, A., Armstrong, B., 2013. Reducing and meta-analysing estimates from distributed lag non-linear models. BMC Med Res Methodol 13, 1. 10.1186/1471-2288-13-1

Gasparrini, A., Armstrong, B., Kenward, M.G., 2012. Multivariate meta-analysis for non-linear and other multi-parameter associations. Stat Med 31, 3821–3839. 10.1002/sim.5471

Gasparrini, A., Guo, Y., Hashizume, M., Lavigne, E., Zanobetti, A., Schwartz, J., Tobias, A., Tong, S., Rocklöv, J., Forsberg, B., Leone, M., De Sario, M., Bell, M.L., Guo, Y.-L.L., Wu, C., Kan, H., Yi, S.-M., De Sousa Zanotti Stagliorio Coelho, M., Saldiva, P.H.N., Honda, Y., Kim, H., Armstrong, B., 2015. Mortality risk attributable to high and low ambient temperature: a multicountry observational study. The Lancet 386, 369–375. 10.1016/S0140-6736(14)62114-0

Godói, I.P., Da Silva, L.V.D., Sarker, A.R., Megiddo, I., Morton, A., Godman, B., Alvarez- Madrazo, S., Bennie, M., Guerra-Junior, A.A., 2018. Economic and epidemiological impact of dengue illness over 16 years from a public health system perspective in Brazil to inform future health policies including the adoption of a dengue vaccine. Expert Review of Vaccines 17, 1123–1133. 10.1080/14760584.2018.1546581

Guzman, M.G., Harris, E., 2015. Dengue. The Lancet 385, 453–465. 10.1016/S0140-6736(14)60572-9

Imai, C., Armstrong, B., Chalabi, Z., Mangtani, P., Hashizume, M., 2015. Time series regression model for infectious disease and weather. Environmental Research 142, 319–327. 10.1016/j.envres.2015.06.040

Jackson, D., Riley, R., White, I.R., 2011. Multivariate meta-analysis: potential and promise. Stat Med 30, 2481–2498. 10.1002/sim.4172

Jacobson, L.D.S.V., Oliveira, B.F.A.D., Schneider, R., Gasparrini, A., Hacon, S.D.S., 2021. Mortality Risk from Respiratory Diseases Due to Non-Optimal Temperature among Brazilian Elderlies. IJERPH 18, 5550. 10.3390/ijerph18115550

Kenney, W.L., Craighead, D.H., Alexander, L.M., 2014. Heat Waves, Aging, and Human Cardiovascular Health. Medicine & Science in Sports & Exercise 46, 1891–1899. 10.1249/MSS.0000000000000325

Kephart, J.L., Sánchez, B.N., Moore, J., Schinasi, L.H., Bakhtsiyarava, M., Ju, Y., Gouveia, N., Caiaffa, W.T., Dronova, I., Arunachalam, S., Diez Roux, A.V., Rodríguez, D.A., 2022. City-level impact of extreme temperatures and mortality in Latin America. Nat Med 28, 1700–1705. 10.1038/s41591-022-01872-6

Lee, S.A., Economou, T., De Castro Catão, R., Barcellos, C., Lowe, R., 2021. The impact of climate suitability, urbanisation, and connectivity on the expansion of dengue in 21st century Brazil. PLoS Negl Trop Dis 15, e0009773. 10.1371/journal.pntd.0009773

Lowe, R., Lee, S.A., O’Reilly, K.M., Brady, O.J., Bastos, L., Carrasco-Escobar, G., De Castro Catão, R., Colón-González, F.J., Barcellos, C., Carvalho, M.S., Blangiardo, M., Rue, H., Gasparrini, A., 2021. Combined effects of hydrometeorological hazards and urbanisation on dengue risk in Brazil: a spatiotemporal modelling study. The Lancet Planetary Health 5, e209–e219. 10.1016/S2542-5196(20)30292-8

Martínez-Solanas, È., Basagaña, X., 2019. Temporal changes in the effects of ambient temperatures on hospital admissions in Spain. PLoS ONE 14, e0218262. 10.1371/journal.pone.0218262

Mistry, M.N., Schneider, R., Masselot, P., Royé, D., Armstrong, B., Kyselý, J., Orru, H., Sera, F., Tong, S., Lavigne, É., Urban, A., Madureira, J., García-León, D., Ibarreta, D., Ciscar, J.-C., Feyen, L., De Schrijver, E., De Sousa Zanotti Stagliorio Coelho, M., Pascal, M., Tobias, A., Multi-Country Multi-City (MCC) Collaborative Research Network, Alahmad, B., Abrutzky, R., Saldiva, P.H.N., Correa, P.M., Orteg, N.V., Kan, H., Osorio, S., Indermitte, E., Jaakkola, J.J.K., Ryti, N., Schneider, A., Huber, V., Katsouyanni, K., Analitis, A., Entezari, A., Mayvaneh, F., Michelozzi, P., de’Donato, F., Hashizume, M., Kim, Y., Diaz, M.H., De La Cruz Valencia, C., Overcenco, A., Houthuijs, D., Ameling, C., Rao, S., Seposo, X., Nunes, B., Holobaca, I.-H., Kim, H., Lee, W., Íñiguez, C., Forsberg, B., Åström, C., Ragettli, M.S., Guo, Y.-L.L., Chen, B.-Y., Colistro, V., Zanobetti, A., Schwartz, J., Dang, T.N., Van Dung, D., Guo, Y., Vicedo-Cabrera, A.M., Gasparrini, A., 2022. Comparison of weather station and climate reanalysis data for modelling temperature-related mortality. Sci Rep 12, 5178. 10.1038/s41598-022-09049-4

Morral-Puigmal, C., Martínez-Solanas, È., Villanueva, C.M., Basagaña, X., 2018. Weather and gastrointestinal disease in Spain: A retrospective time series regression study. Environment International 121, 649–657. 10.1016/j.envint.2018.10.003

Muñoz-Sabater, J., Dutra, E., Agustí-Panareda, A., Albergel, C., Arduini, G., Balsamo, G., Boussetta, S., Choulga, M., Harrigan, S., Hersbach, H., Martens, B., Miralles, D.G., Piles, M., Rodríguez-Fernández, N.J., Zsoter, E., Buontempo, C., Thépaut, J.-N., 2021. ERA5- Land: a state-of-the-art global reanalysis dataset for land applications. Earth Syst. Sci. Data 13, 4349–4383. 10.5194/essd-13-4349-2021

National Institute of Meteorology (INMET), 2024. Metheorological database, INMET. [WWW Document]. URL https://bdmep.inmet.gov.br/# (accessed 2.19.24).

Noronha, K.V.M.D.S., Guedes, G.R., Turra, C.M., Andrade, M.V., Botega, L., Nogueira, D., Calazans, J.A., Carvalho, L., Servo, L., Ferreira, M.F., 2020. Pandemia por COVID-19 no Brasil: análise da demanda e da oferta de leitos hospitalares e equipamentos de ventilação assistida segundo diferentes cenários. Cad. Saúde Pública 36, e00115320. 10.1590/0102-311x00115320

Paz-Bailey, G., Adams, L.E., Deen, J., Anderson, K.B., Katzelnick, L.C., 2024. Dengue. The Lancet 403, 667–682. 10.1016/S0140-6736(23)02576-X

Pudpong, N., Hajat, S., 2011. High temperature effects on out-patient visits and hospital admissions in Chiang Mai, Thailand. Science of The Total Environment 409, 5260–5267. 10.1016/j.scitotenv.2011.09.005

Royé, D., Íñiguez, C., Tobías, A., 2020. Comparison of temperature–mortality associations using observed weather station and reanalysis data in 52 Spanish cities. Environmental Research 183, 109237. 10.1016/j.envres.2020.109237

Saha, M.V., Davis, R.E., Hondula, D.M., 2014. Mortality Displacement as a Function of Heat Event Strength in 7 US Cities. American Journal of Epidemiology 179, 467–474. 10.1093/aje/kwt264

Schneider, A., Rückerl, R., Breitner, S., Wolf, K., Peters, A., 2017. Thermal Control, Weather, and Aging. Curr Envir Health Rpt 4, 21–29. 10.1007/s40572-017-0129-0

Schwartz, J., Samet, J.M., Patz, J.A., 2004. Hospital Admissions for Heart Disease: The Effects of Temperature and Humidity. Epidemiology 15, 755–761. 10.1097/01.ede.0000134875.15919.0f

Silveira, I.H., Oliveira, B.F.A., Cortes, T.R., Junger, W.L., 2019. The effect of ambient temperature on cardiovascular mortality in 27 Brazilian cities. Science of The Total Environment 691, 996–1004. 10.1016/j.scitotenv.2019.06.493

Suaya, J.A., Shepard, D.S., Siqueira, J.B., Martelli, C.T., Lum, L.C.S., Tan, L.H., Kongsin, S., Jiamton, S., Garrido, F., Montoya, R., Armien, B., Huy, R., Castillo, L., Caram, M., Sah, B.K., Sughayyar, R., Tyo, K.R., Halstead, S.B., 2009. Cost of dengue cases in eight countries in the Americas and Asia: a prospective study. Am J Trop Med Hyg 80, 846– 855.

Vaidyanathan, A., Saha, S., Vicedo-Cabrera, A.M., Gasparrini, A., Abdurehman, N., Jordan, R., Hawkins, M., Hess, J., Elixhauser, A., 2019. Assessment of extreme heat and hospitalizations to inform early warning systems. Proc. Natl. Acad. Sci. U.S.A. 116, 5420–5427. 10.1073/pnas.1806393116

Van Wyk, H., Eisenberg, J.N.S., Brouwer, A.F., 2023. Long-term projections of the impacts of warming temperatures on Zika and dengue risk in four Brazilian cities using a temperature-dependent basic reproduction number. PLoS Negl Trop Dis 17, e0010839. 10.1371/journal.pntd.0010839

Wang, B., Chai, G., Sha, Y., Zha, Q., Su, Y., Gao, Y., 2021. Impact of ambient temperature on cardiovascular disease hospital admissions in farmers in China’s Western suburbs. Science of The Total Environment 761, 143254. 10.1016/j.scitotenv.2020.143254

Werneck, G.L., Macias, A.E., Mascarenas, C., Coudeville, L., Morley, D., Recamier, V., Guergova-Kuras, M., Puentes-Rosas, E., Baurin, N., Toh, M.-L., 2018. Comorbidities increase in-hospital mortality in dengue patients in Brazil. Mem. Inst. Oswaldo Cruz 113. 10.1590/0074-02760180082

Wibawa, B.S.S., Wang, Y.-C., Andhikaputra, G., Lin, Y.-K., Hsieh, L.-H.C., Tsai, K.-H., 2024. The impact of climate variability on dengue fever risk in central java, Indonesia. Climate Services 33, 100433. 10.1016/j.cliser.2023.100433

Yi, X., Chang, Z., Zhao, X., Ma, Y., Liu, F., Xiao, X., 2020. The temporal characteristics of the lag-response relationship and related key time points between ambient temperature and hand, foot and mouth disease: A multicity study from mainland China. Science of The Total Environment 749, 141679. 10.1016/j.scitotenv.2020.141679

Zhao, Q., Li, S., Coelho, M.S.Z.S., Saldiva, P.H.N., Hu, K., Huxley, R.R., Abramson, M.J., Guo, Y., 2019. The association between heatwaves and risk of hospitalization in Brazil: A nationwide time series study between 2000 and 2015. PLoS Med 16, e1002753. 10.1371/journal.pmed.1002753

